# Intent Drift in LLM-Assisted BCI Communication: An In-Silico Benchmark Under Simulated Decoder Corruption

**DOI:** 10.64898/2026.08.20.26360939

**Authors:** Alon Gorenshtein, Mahmud Omar, Eric Jia, Yosef Adiniaev, Oved Daniel, Jonathan Kruskal, Muneeb Ahmed, Olga Brook, Eyal Klang, Yiftach Barash

## Abstract

**Background:** Large language models (LLMs) are increasingly used to correct noisy, error-prone text typed through brain-computer interfaces (BCIs) and other assistive communication devices. A fluent correction can still be wrong: the model can substitute a different intended message, a failure we call intent drift. Whether meaning survives correction, and whether model confidence flags failure, is unmeasured.

**Methods:** We built an in-silico benchmark: 20 open-weight LLMs corrected simulated P300 speller text under five levels of decoder error (0-40% character error rate). Testing used the full ALS message-banking vocabulary, a set of clinically important messages (eg, involving a medication dose), and matched controls. Each of 4,252,326 outputs was scored faithful, degraded, or drifted by an automated system benchmarked against physicians. A separate subanalysis compared six correction strategies across seven models.

**Findings:** Drift rose steeply with decoder error, from 2.2% at no error to 60.3% at the most severe level tested (odds ratio 2.30 per 10-percentage-point increase)-a stress-test ceiling, not an expected real-world rate. Model confidence distinguished correct from incorrect outputs reasonably well (AUROC 0.83) but overstated its own reliability: 28.4% of high-confidence outputs were not faithful. Clinically important messages drifted slightly more than matched controls (odds ratio 1.10). At low error rates, corrections succeeded more often than they went confidently wrong, but this reversed above 20-30% error. No correction strategy eliminated drift: cautious approaches reduced it, permissive ones increased it, and even the best still produced a wrong message in 18 of 100 corrections. Physician review of a sample agreed moderately with automated scoring (kappa 0.41); adjusting for this disagreement lowered but did not remove the pattern of rising drift with decoder error (31.4% to 28.3%).

**Interpretation:** LLMs correcting BCI text produced fluent but sometimes wrong messages, more often as decoding quality worsened, and their own confidence did not reliably warn when this happened. These in-silico findings support evaluating such systems for meaning preservation, not only speed and accuracy, before deployment; prospective, human-in-the-loop evaluation is needed.

**Funding:** Harvard Catalyst (CTSA UL1TR002541).

**Research in context:** *Evidence before this study:* We searched PubMed, arXiv, and bioRxiv from database inception to July 19, 2026, combining terms for large language models with brain-computer interfaces, P300 spellers, and augmentative and alternative communication, without language restriction. Exact strings and record counts are in eMethods S9. Prior studies integrating large language models with P300 spellers and communication BCIs reported keystroke savings, typing speed, information transfer rate, and character-level accuracy. Some concluded that correction was near-optimal once residual errors were manually fixed, placing meaning-level change outside their frame. No study had measured how often post-editing changed a message’s intent, whether this depended on decoder corruption, or whether stated confidence tracked whether the output was faithful.

*Added value of this study:* This in-silico benchmark measures intent drift and confidence calibration in language-model post-editing as a function of decoder corruption, reported separately across AUTH, a message-critical set, and matched controls. Using an empirical P300 confusion matrix and 4,252,326 labeled generations across 20 open-weight models, the primary benchmark questions found detected drift rose steeply with corruption and stated confidence discriminated faithful outputs well but was poorly calibrated; secondary analyses found message-critical content carried excess drift that survived detector removal and that detected drift varied more than two-fold across models (20.8-49.3% in AUTH). A separate subanalysis of six correction strategies (original seven-model panel) found no strategy removed drift: conservative editing or abstention lowered it, while alternatives or expansion raised it.

*Implications of all the available evidence:* Evaluations of language-model-assisted communication BCIs and AAC systems should report intent drift and confidence calibration alongside speed and accuracy, should test message-critical content separately from routine messages, and should treat interface policy as a measured design variable. Because the construct is communicative intent, the absence of patient and public involvement in probe-set design is material. Because confidence did not reliably flag drift, it alone cannot gate human review. These are in-silico findings; they support the need for prospective, human-in-the-loop evaluation before deployment implications can be drawn.

## Introduction

Communication brain-computer interfaces (BCIs) restore a channel for expression to people who have lost speech and movement to severe paralysis, including amyotrophic lateral sclerosis (ALS) and brainstem stroke.^1,2^ Systems such as the P300 speller let a user select characters or words from decoded neural signals, but the resulting stream is slow and error-prone.^3^ Large language models (LLMs) are increasingly proposed at the interface layer of these systems,^4–7^ and of augmentative-and-alternative-communication (AAC) systems more broadly,^8,9^ to post-edit noisy decoded text into fluent, intended messages.

A language model fluent enough to reconstruct a noisy character stream into a well-formed message can also reconstruct it into one that asserts a different intent than the user meant to send. A garbled request for water can be completed as a fluent request for a blanket: both are plausible surface completions of the corrupted input, and only the sender knows which was meant. This failure is distinct from decoding accuracy: the output can be grammatical and assertive while substituting one plausible message for another, an assumption speed and character-accuracy metrics do not capture. We refer to this failure mode as semantic intent substitution, or informally, intent drift. A separate question is calibration: whether a model signals its own uncertainty when a reconstruction drifts, or returns the same confident tone either way.

Evaluations of LLM-assisted spellers have measured keystroke savings, typing speed, information transfer rate, and character-level correction accuracy.^4–7^ Two recent reports come closest: one reconstructing single sentences from a P300 speller, one treating language modeling as solved after manually correcting residual errors before scoring (Discussion).^5–7^ Neither varied decoder corruption systematically, measured how often a reconstruction asserted an unintended message, or tested whether confidence tracked that risk. The stakes are concrete for the people who depend on these systems: AAC users have reported apprehension that automated suggestions will not reflect what they meant to say.^10^ Our own systematic review of LLM-BCI communication systems named this failure mode as a conceptual concern but did not measure it.^4^

In this study, we built an in-silico benchmark of the semantic fidelity of LLM-assisted post-editing. Corruption was applied synthetically, calibrated to an empirical P300-speller confusion matrix, so decoder error could be varied in a controlled, reproducible way. Using an ALS message-banking vocabulary (AUTH) and a message-critical challenge set with matched controls, we measured, across 20 open-weight LLMs, the detected drift rate as a function of decoder corruption and the calibration of each model’s stated confidence against its correctness. The two primary benchmark questions were whether drift rises with corruption in a dose-dependent way and whether models signal, through confidence, when a reconstruction is wrong.

## Methods

### Study design and reporting

This was an in-silico controlled benchmark of language-model post-editing on simulated P300-speller output; no participant used a brain-computer interface, and no clinical or device-use claim is made. Reporting followed TRIPOD-LLM^11,12^ and the LLM-BCI reporting checklist^4^ (eTable 1), adapted to a simulation rather than a patient-facing design.

### Message corpora

Three message sets were used (Results; eMethods S1). AUTH was the full Costello/Boston Children’s Hospital ALS message-banking vocabulary,^13^ all 2,168 phrases across 40 categories, analyzed as a census (eMethods S1). CRIT was 131 message-critical items frozen before the main run, spanning five categories where a reconstruction error changes clinically consequential content: negation, numeral or dose, recipient, urgency, and actionable omission. Each CRIT item was matched 1:1 to a control (CTRL, n=131) by greedy nearest-neighbor matching on five covariates (eMethods S3). CTRL is a 131-message subset of AUTH; CRIT is fully disjoint from both, so the pooled panel spans 2,299 distinct messages, not 2,430.

### Corruption model (exposure)

Corruption was synthetic, not output from a live decoding session, parameterized by an empirical confusion matrix from P300-speller selections by ALS participants in bigP3BCI studies F, L, and N,^14^ pooled on the canonical 6-by-6, 36-symbol BCI2000 grid^15^ (eTable 13). Two corruption-model assumptions were not empirically validated (Limitations; eTable 13). Each message was corrupted at five target character-error rates (0, 0.1, 0.2, 0.3, 0.4), 20 replicate corruptions per message-by-target-rate cell from deterministic seeds. Target CER is the primary exposure throughout; the realized outcome (true Levenshtein CER, error count, corrupted-negation/numeral flags) is a secondary exposure in every downstream model. This gave 243,000 exposures for 17 of 20 models; the remaining three (GPT-OSS-20B, GPT-OSS-120B, and DeepSeek-R1-32B) used a reduced-replicate grid of up to 40,500 each for their added chain-of-thought inference cost^16^ (eMethods S1); GPT-OSS-20B reached 40,326 (174 generations failed or were unparseable and excluded), for 4,252,326 labeled generations across the full panel.

### Models and prompting condition

Twenty open-weight instruction-tuned models were served at temperature 0, identified throughout by their served tag (full list, digests, and quantizations in eTable 14); the three reasoning models also received a larger `output-token limit` for their chain-of-thought trace. They were served at differing quantizations^17^ (Discussion, Limitations). All models were queried with a single frozen post-edit prompt (P0: reconstruct the intended message from the noisy character stream without adding content), one of a six-prompt bank frozen before the main run; the other five prompts define the interface-condition subanalysis described next.

### Interface-condition subanalysis

The main analysis holds the interface policy fixed at forced reconstruction (always returns a single reconstruction; no abstention, hedging, or alternatives). The remaining five prompts define the interface conditions: minimal edit (P1, correct spelling only), copy when uncertain (P2, change as little as possible), abstention enabled (P3, decline rather than guess), candidate list (P4, one best reconstruction plus up to two alternatives), and expansion (P5, expand the terse stream into the full intended sentence), re-run on a 562-message subset (300 AUTH, 131 CRIT, 131 matched CTRL) across all five corruption levels, 10 replicates, and the original seven-model panel (196,700 generations per arm; Discussion, Limitations; eMethods S8).

Abstention sits outside the faithful, degraded, and drift taxonomy: declines were counted separately, never entering the drift denominator; the subanalysis also reports faithful/drifted messages per 100 exposures attempted, counting declines. Condition effects on drift used message-clustered cluster-robust logistic regression against forced reconstruction, averaged over the corruption grid; corruption-resolved rates are also reported directly (eMethods S8).

### Outcome taxonomy

Each reconstruction received one of three labels, faithful, degraded, or drift, plus an independent fluency flag: fluent meant the output read as ordinary, well-formed language regardless of correctness; a faithful output was fluent and conveyed the intended message; a degraded output was disfluent or incomplete and asserted no different meaning; a drift output was fluent but conveyed a different intent. Meaning difference was adjudicated by a bidirectional natural-language-inference contradiction check, a cosine-similarity threshold, and five rule-based detectors matched to the CRIT categories (eMethods S4). Message-critical status is a fixed input property assigned before corruption, not an outcome label; any message-critical item could resolve as faithful, degraded, or drift.

### Automated evaluation

Outcome labels came from a multi-evaluator ensemble, two natural-language-inference checkpoints^18^ and two sentence-embedding checkpoints^19^ alongside a shared fluency model, disagreement resolved by a tie-break rule (eMethods S5). This pipeline’s labels were characterized against a blinded physician panel, with that comparison and a misclassification-corrected sensitivity analysis in Results and eMethods S6.

### Statistical analysis

The primary target-CER dose-response was a binary logistic regression with target CER as a continuous ordered exposure, message-only and message-by-model two-way cluster-robust, with per-model slopes (eTable 3). Confidence calibration, the second primary endpoint, was summarized per model by the expected calibration error^20^ and the AUROC for verbalized confidence^21,22^ predicting a faithful output, message-clustered bootstrap intervals, pooled across models by DerSimonian-Laird random-effects meta-analysis^23^ (eTables 11-12; Discussion, Limitations).

Realized corruption was analyzed as a secondary dose-response via message-clustered, cluster-robust logistic regression on realized corruption and message-level covariates (Limitations; eMethods S7). Excess drift from message-critical content used a matched-pair conditional logistic regression conditioned on the CRIT-CTRL pair at identical realized corruption (eMethods S7). A Rogan-Gladen^24^ misclassification correction is reported from the physician-panel-measured ensemble error rates (eMethods S7); sensitivity analyses varied the fluency threshold and tie-break rule. Pooled drift rates are generation-weighted; an equal-model-weight sensitivity check is also reported (Results; eTable 2).

## Results

### Sample

The benchmark yielded 4,252,326 labeled generations from 20 open models, spanning 2,299 distinct messages across AUTH, CRIT, and CTRL: 3,793,828 AUTH, 229,249 CRIT, and 229,249 matched CTRL exposures. The interface-condition subanalysis remained on the original seven-model panel; physician-panel validation centers on an earlier 16-model panel that predates the four models added last (below; Discussion, Limitations).

Because AUTH and CRIT draw from different message populations, and CTRL rather than AUTH is the matched comparator for CRIT, results are reported by corpus rather than pooled. Across the expanded grid, detected drift was 31.5% in AUTH, 33.7% in CRIT, and 31.5% in CTRL (faithful/degraded rates in eTable 3), averaged over the five-level 0-40% target-CER stress grid, so these are stress-test summaries, not expected clinical event rates (Figure 1a; Figure 2). These pooled rates are generation-weighted (Methods); equal-model-weighting gave similar results (eTable 2).

**Figure 1.**
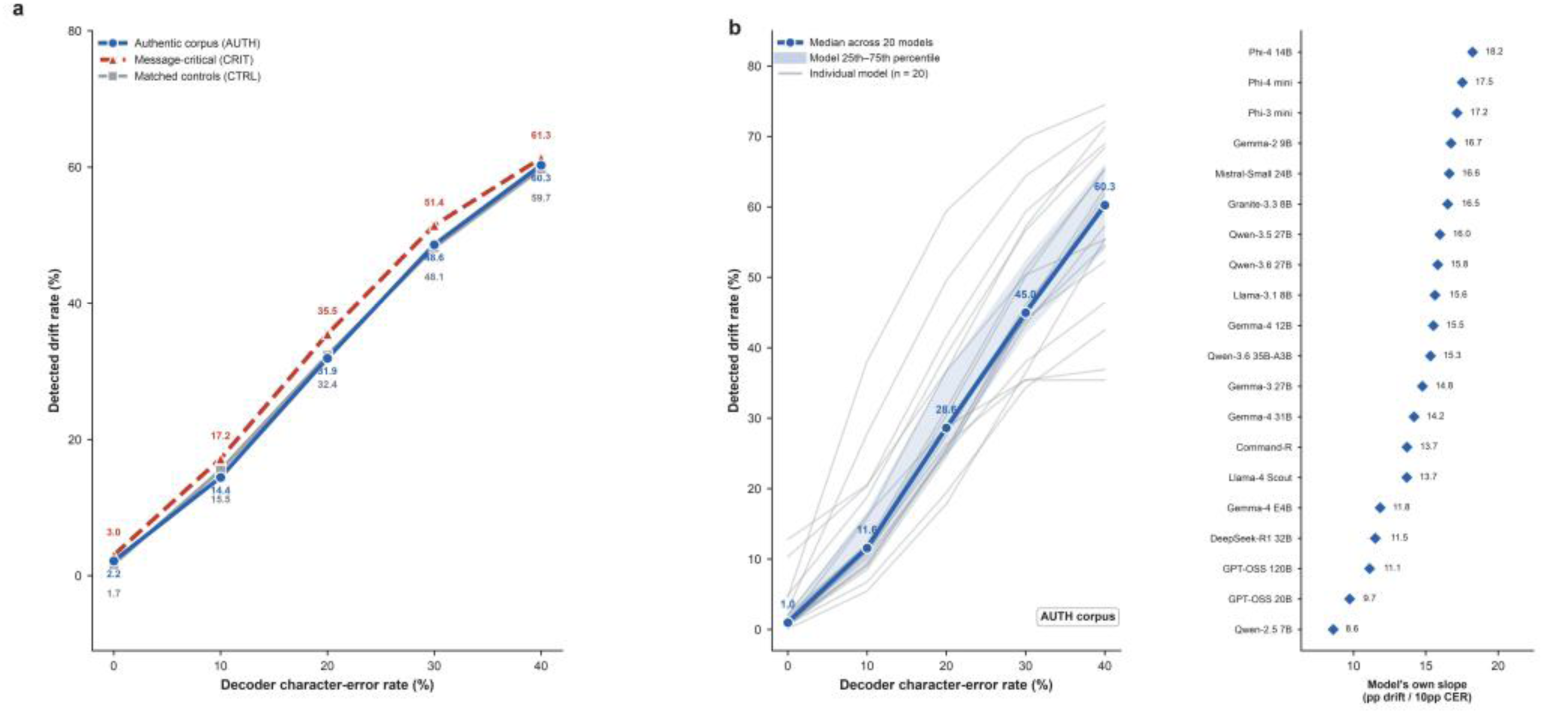
Detected drift by corpus and by model, across in-silico decoder corruption. (a) Detected drift rate as a function of decoder character-error rate (CER), plotted separately for the ALS message-banking vocabulary (AUTH), the message-critical probe set (CRIT), and matched controls (CTRL). Each point is the percentage of evaluated generations scored drift at that CER level (0-40%), with the value printed on the point; vertical caps are Wilson-score 95% CIs (narrower than the marker for AUTH and CTRL, whose per-cell counts are large). (b) Model heterogeneity in detected drift versus CER, AUTH corpus. Left: each of the 20 evaluated models’ own drift-versus-CER curve, drawn as a faint individual line with no per-model color or legend, overlaid with the bold across-model median (value printed at each CER level) and a shaded 25th-75th-percentile band. Right: each model’s own least-squares slope (percentage points of drift per 10-point rise in CER), one dot per model, ordered ascending, with a message-clustered bootstrap 95% CI (1,000 resamples of the authentic-corpus messages) drawn through each dot. The models differ in checkpoint and quantization, so this panel shows available open models stress-tested identically, not a controlled like-for-like architecture sweep; the full 20-model hardware and digest manifest is in eTable 14.

**Figure 2.**
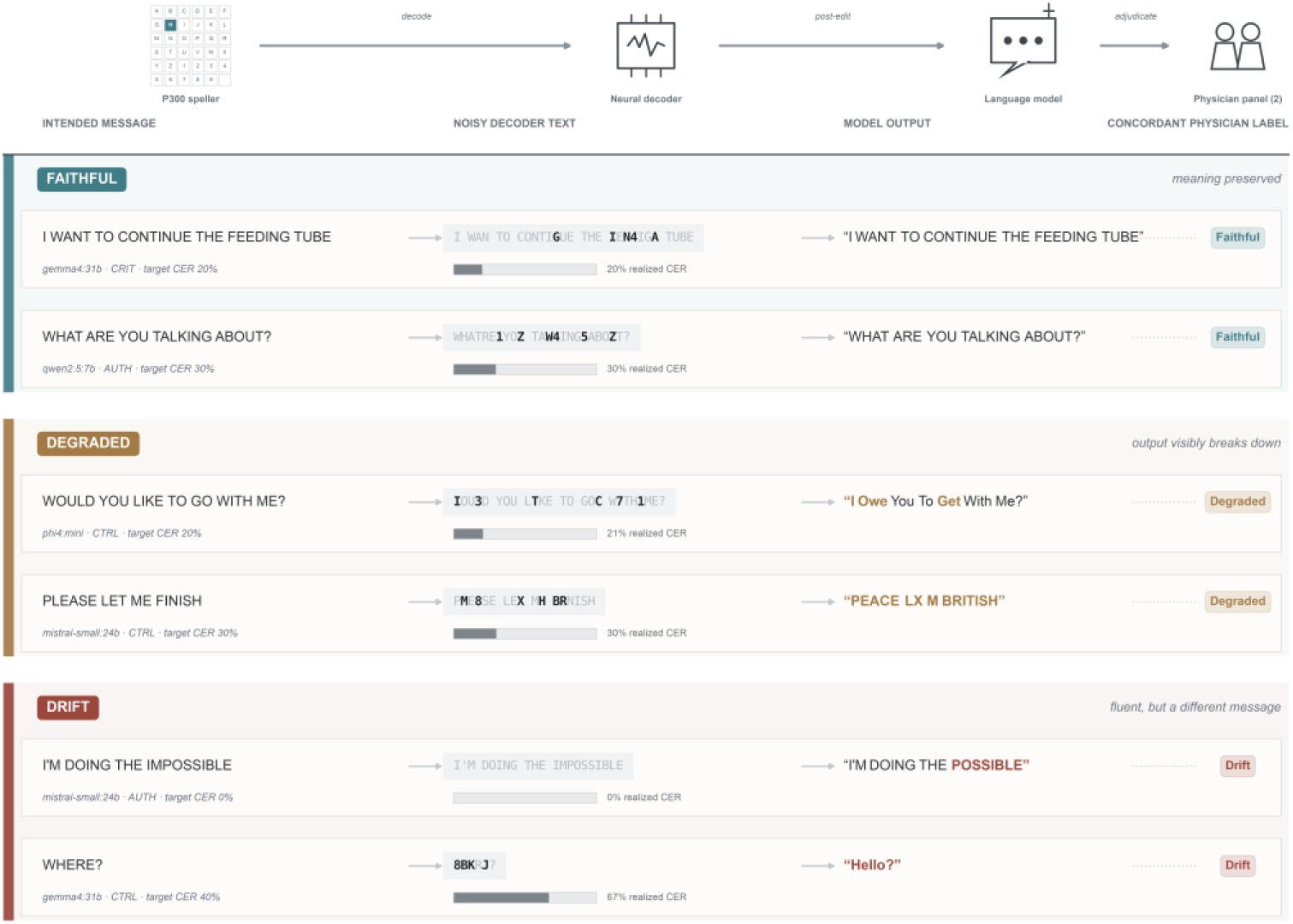
Physician-validated faithful, degraded, and drift reconstruction examples. The schematic (top) mirrors the columns below: a message attempted on a P300 speller is turned into noisy text by the neural decoder, post-edited by a language model, and adjudicated by a two-physician panel into the concordant label. Six example generations follow (two per outcome class), shown verbatim from the labeled attempt table with no text edited for display. Every row’s three-class outcome (faithful, degraded, or drift) is one on which both physician raters of the completed 16-model panel (eMethods S6), blinded to model identity and corruption level, independently gave the same label; no automated-only, single-rater, or adjudicator-only label is shown, and no unvalidated subtype (for example, a negation flip or a numeral change) is captioned. In the noisy decoder text, the characters that differ from the intended message (the corruption the decoder introduced) are set in bold monospace against the unchanged characters; in the model output, the words the model altered relative to the intended message are set in the outcome colour and the preserved words in grey, so a drift row shows the specific changed word and a faithful row reads as intact. Faithful and degraded rows are drawn at two moderate, target-aligned corruption levels (20% and 30% target character error rate); drift rows span the realized range, one at essentially no decoder corruption and one at the highest, showing that a fluent, confidently wrong reconstruction can occur both with and without measurable corruption. The bar beneath each decoder cell is that row’s realized character error rate; the model, corpus, and target character error rate are printed under each row.

### Drift rose steeply with decoder corruption

Detected drift rose monotonically with CER in all corpora (Figure 1a; eTable 3). In AUTH, it increased from 2.2% at 0% CER to 60.3% at 40%; CRIT and CTRL showed similar gradients. Each 10-percentage-point increase more than doubled the odds of drift (odds ratio 2.30, 95% CI 2.27-2.32 with message clustering; 2.15-2.45 with message-by-model clustering), with positive slopes in every model (5.14-10.55 log-odds; median 8.45). Realized CER showed the same direction in secondary analysis (odds ratio 1.61 per 10 percentage points; log-odds 4.77, 95% CI 4.43-5.11; eTable 12).

### Faithful rescues exceeded fluent errors at low but not high corruption

Relative to sending the raw decoded stream, and holding the interface policy fixed at forced reconstruction (Methods; alternative policies below), the model turned a visibly degraded decode into a faithful message in 26.7% of transitions (rescue) and into a fluent but different message in 29.7% (silent failure); drift from an already-faithful raw stream was 0.8%. The net faithful messages gained per fluent error introduced (an unweighted count) was 0.88 overall but fell steeply with corruption, from 3.37 at 10% target CER to 0.29 at 40%, dropping below 1 above roughly 20 to 30% CER as a heavily corrupted input no longer constrained the reconstruction.

### No interface policy removed drift

Relative to forced reconstruction (drift 31.5%; Methods), detected drift was lower under copy-when-uncertain, minimal edit, and permitted abstention (odds ratios 0.75-0.85) and higher under candidate list and expansion (1.13 and 1.25), averaged over the corruption grid, all 95% CIs excluding 1 (eTable 7).

Abstention was targeted rather than indiscriminate: the decline rate rose from 5.6% at no target corruption to 52.2% at 40%, yet it still answered 47.8% of the most corrupted inputs and drifted on 57.5% of those answers. Counting declines directly, permitted abstention returned 18.0 drifted messages per 100 exposures against 31.5 under forced reconstruction, at a cost of 6.8 faithful messages (49.9 versus 56.7; eTable 7, Panel B).

The benefit-harm ranking followed the same order (unweighted count; eTable 7): faithful messages gained per fluent error introduced ranged from 1.35 (permitted abstention) to 0.83 (expansion), with forced reconstruction at 0.92 in this 562-message subanalysis (versus 0.88 across the full dataset above). A faithful reconstruction appeared somewhere in the candidate list for 59.0% of exposures against 55.0% for the primary answer alone (eTable 7, Panel D). No condition brought drift near zero.

### Confidence discriminated faithful outputs well but was poorly calibrated

Stated confidence discriminated faithful outputs better than it flagged drift specifically. The meta-analytic AUROC(confidence→faithful) was 0.83 (95% CI, 0.80-0.85), while the expected calibration error (ECE) was 0.32 (95% CI, 0.27-0.37), so confidence systematically overstated the probability of a faithful output (Figure 3, eFigure 4); 15,814 outputs with missing verbalized confidence were dropped. At confidence 90 or higher, 28.4% were not faithful, varying widely across the panel (5.4% to 64.0%; eTable 2). Missing confidence was itself informative, enriched roughly two-fold for drift (67.9% versus a 31.6% baseline among outputs with valid confidence; eTable 11).

**Figure 3.**
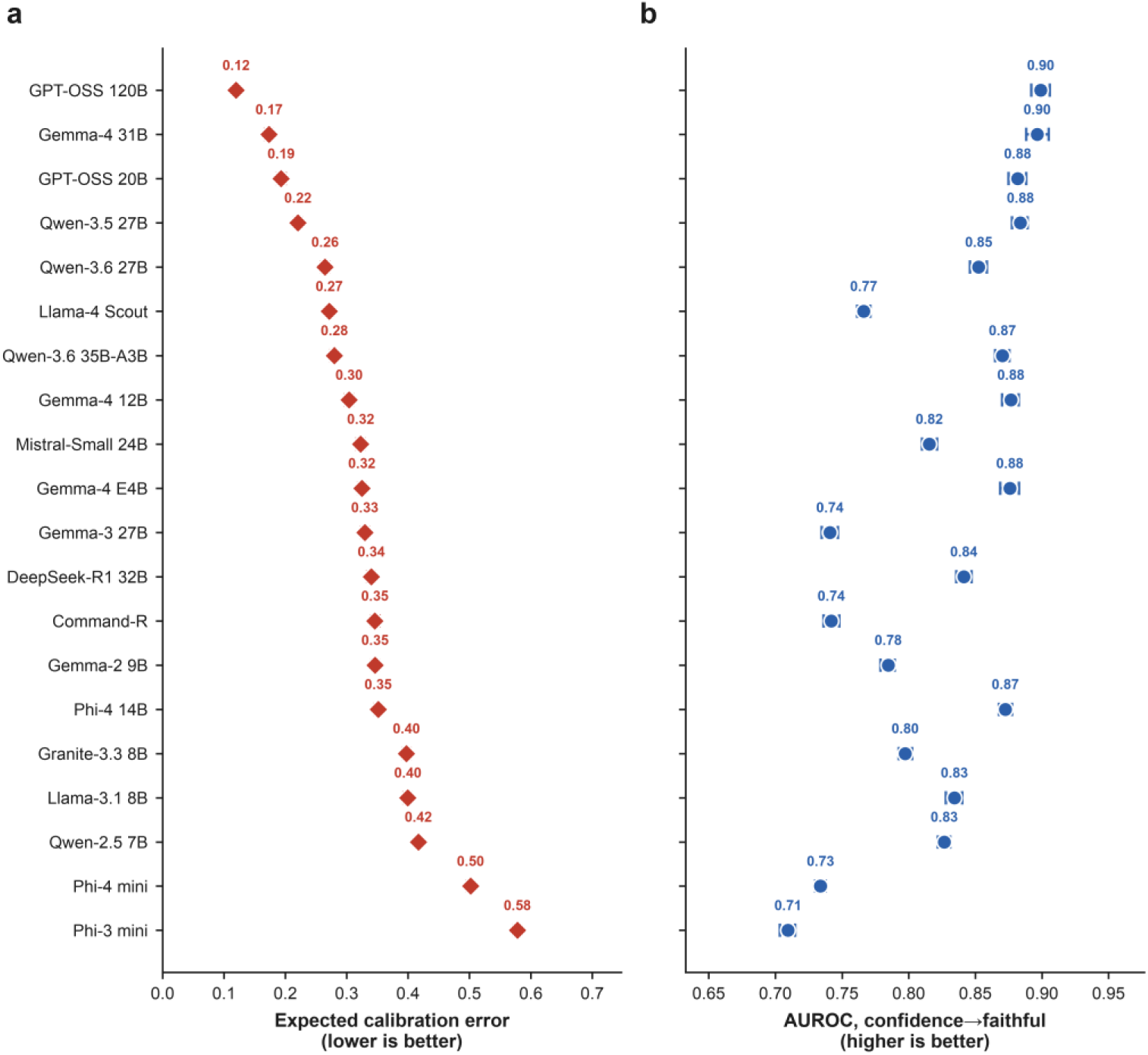
Per-model confidence calibration: expected calibration error and discrimination. (a) Expected calibration error (ECE, lower is better) for stated confidence discriminating faithful from non-faithful (degraded or drift) outputs, one point per evaluated model, sorted by ECE; faithful was coded 1 and degraded and drift were coded 0 on the same outputs, and values are printed at each point. Horizontal caps are each model’s 95% CI (message-clustered; each model contributes roughly 2,300 messages, so the ECE intervals are narrower than the marker). (b) AUROC for the same discrimination task (higher is better), the 20 models in the same row order as (a), each with its message-clustered 95% CI. ECE and AUROC are distinct metrics with opposite preferred directions and are shown as two separate aligned dot plots, not joined by a connecting line. Every model was substantially overconfident (ECE well above 0) while retaining real ranking ability (AUROC well above 0.5): confidence discriminated faithful outputs but overstated their probability. Verbalized confidence is not on a common scale across models (Methods), so per-model calibration, not a single pooled curve, is the interpretable display. The pooled confidence-outcome curve and the pooled meta-analytic ECE (0.32, 95% CI 0.27-0.37) and AUROC (0.83, 95% CI 0.80-0.85), reported as a descriptive cross-model summary, are in eFigure 4, and per-model reliability curves in eFigure 3.

A blinded 148-item check of automated zero-realized-CER drift labels (broader than target- 0% in Figure 1; 22,010 total) found only 29.7% physician-confirmed (kappa 0.752; eTable 6, Panel E).

### Challenge messages carried excess drift after matching

In the original seven-model panel, the raw CRIT-CTRL gap persisted after matching (eTable 4). Because the five detectors flagging a critical substitution also define the CRIT categories, this could be partly circular; the rule-free re-estimate, drift decided by the meaning channel alone (Methods), is the primary criticality contrast: OR 1.10 (95% CI, 1.07-1.13; eTable 9), a matched absolute risk difference of 2.2 percentage points (95% CI, 0.4-4.2), small and not a clean gradient across corruption levels. A 20-model extension gave a similar excess (OR 1.10, 95% CI 1.08-1.11; eTable 9).

### Sensitivity analyses

The dose-response slopes were insensitive to deduplication (under 4%; eTable 8); detected drift shifted more under alternative tie-break rules and fluency thresholds (27.6-34.7% versus 31.0% primary; eTable 5).

### Automated labels agreed with the physician panel on the faithful and drift classes

Two physicians blindly rated 2,281 items spanning the 16 models (eMethods S1), 3 corpora, the full CER grid, and all three outcome classes; a third (O.D.) adjudicated the 343 disagreements. Inter-rater kappa was 0.759; ensemble-versus-consensus kappa was 0.412 (moderate), class F1 high for faithful and drift, low for degraded, most often relabeled as drift, concordant with an earlier seven-model subset (eTable 6, Panels A-E).

### A sampling-weighted misclassification correction, applied to the 16-model panel, lowered rather than raised detected drift

Using the physician-measured confusion matrix and a message-clustered bootstrap, the Rogan-Gladen correction moved pooled drift from 31.4% to 28.3%, AUTH from 31.3% to 27.9% (95% CI, 22.1-33.0), CRIT from 33.5% to 33.4%, and CTRL from 31.3% to 29.6%. The seven-model sensitivity subset moved the opposite way, from 30.9% to 36.4% (95% CI, 24.5-47.1) in AUTH, with wide, overlapping intervals (eTable 6, Panel D); we treat the 16-model direction as primary, the larger and more recent physician-rated sample (Discussion, Limitations).

### The physician-consensus-corrected dose-response rises with corruption at the same five target-CER levels as the automated one

Applying the weighted Rogan-Gladen correction separately at each of the five target-CER levels, rather than one correction pooled across levels, physician-consensus-corrected drift in the 16-model, three-corpus-pooled panel was 0.0% at 0% target CER (95% CI, 0.0-96.8; a wide interval reflecting near-zero true prevalence rather than a precise null), 12.2% at 10%, 36.2% at 20%, 43.4% at 30%, and 49.3% at 40% (eTable 6, Panel F); the human-consensus-corrected rate rose monotonically with target CER. This rules out a single pooled correction masking a stratum-specific error pattern: the zero-realized-CER physician check above shows exactly that differential error at the lowest end, so the primary dose-response is validated at each level, not only in aggregate.

### Detected drift varied more than two-fold across models

Within AUTH, detected drift ranged from 20.8% (GPT-OSS-120B) to 49.3% (Phi-3-mini) across the 20-model panel (Figure 1b; eTable 2), descriptive variation across checkpoints-as-served since checkpoint, quantization, and serving configuration are confounded with model identity (three models are reduced-replicate chain-of-thought reasoning models; eMethods S1; Discussion, Limitations).

## Discussion

In this in-silico benchmark of 4,252,326 language-model reconstructions, detected semantic drift rose steeply with decoder corruption in all three corpora. The models remained poorly calibrated when they drifted, so high stated confidence was not a sufficient gate. Message-critical content carried a small excess of drift, present in both the seven- and 20-model panels, that survived removing the rule detectors that could have made the contrast circular. Under forced reconstruction, corrections succeeded more often than they went confidently wrong at low corruption, but this reversed above roughly 20 to 30% character-error rate; across six correction strategies, editing conservatively or declining lowered drift while content-adding designs raised it, but none removed it.

When a language model post-edits at the interface layer and the input is too degraded to constrain the reconstruction, it supplies the missing content from its prior, and that content can differ from the intended message while still reading as ordinary language. We use intended-message semantic substitution as an occasional synonym for this pattern to keep the construct precise: fidelity to a known source phrase the sender intended, not access to private subjective intent, which this design cannot observe. That is what separates drift from a degraded output: a degraded output gives a reader a visible cue to distrust it, whereas a drifted output is grammatical and carries no such cue. This is an in-silico measurement of model behavior on a fixed exposure grid, not a claim that a communicative error reached or affected a person (Limitations).

Two recent reports are the closest comparison, and both measured throughput rather than semantic fidelity or calibration: a demonstration connecting a P300 speller to a language model reconstructed intended sentences without an aggregate distribution of meaning-level errors,^5^ and a performance-bounds analysis treated language modeling as solved after correcting residual errors, so meaning-level substitutions lay outside its frame.^6,7^ The AAC literature sets the stakes: users have reported concern automated suggestions would not represent what they meant,^10^ over-prediction could insert unintended words,^25^ and models introduce variations under uncertain input.^26^ Electroencephalogram-to-text decoding similarly yields fluent output only loosely grounded in the neural signal,^27,28^ a pattern consistent with the broader hallucination literature in language generation.^29^ Our earlier systematic review named intent drift as a plausible failure mode but did not measure it.^4^

A benchmark of this form lets an interface model be selected and monitored on semantic fidelity and calibration, not decoding speed and accuracy alone: detected drift varied more than two-fold across the panel, and because high-confidence outputs were not always faithful, a confidence-gated rule would pass some of the substitutions it was meant to catch. The matched excess on message-critical content indicates typical-case and consequential-content fidelity are distinct axes an evaluation reporting only the first would miss. Content-adding interface policies moved fidelity the wrong way, and the policy that cut drift most did so by declining half the most corrupted inputs, a cost a user rather than a benchmark would accept.

### Limitations

This benchmark was in-silico: no person used a brain-computer interface, the corruption was synthetic, and the messages were researcher-assembled; whether drift occurs at this rate in deployment is unknown.

Two of the corruption model’s parameters were not empirically validated, the indel split and the independence of character errors across message positions, so the exposure should be read as P300-informed rather than P300-reproducing (Methods; eTable 13).

The automated labeling pipeline was evaluated against physician judgment rather than validated by it: ensemble-versus-consensus agreement in the 16-model panel was only moderate (kappa 0.41), capturing faithful and drift well but degraded poorly, and a misclassification-corrected estimate moved in opposite directions across panels with wide intervals on the smaller one. Even so, the physician-consensus-corrected rate still rose with corruption at each target-CER level (Results; eTable 6, Panel F); agreement on whether a drift was message-critical was poor. This panel is not external: its physicians, including the corresponding author, contributed to the study’s conceptualisation, methodology, software, and formal analysis, so it is best read as a blinded author-rater panel. Clinician judgment is also an imperfect proxy for communicative intent, and the absence of patient and public involvement in probe-set design is a limitation a qualitative CRIT-taxonomy review would address. The panel predates four models added to reach the full 20-model panel, whose labels carry the same unvalidated status without a direct physician check.

The realized-corruption dose-response was specified as a crossed random-intercept mixed model; its variational-Bayes fit did not converge at full scale, so the reported estimate is a cluster-robust logistic regression without variance components (eMethods S7).

The matched CRIT-CTRL excess (eTable 4, eTable 9) rests on 131 pairs, one a far outlier on match distance; refitting with that pair excluded was numerically singular, so whether the excess holds without it is unreported (eTable 4/9, Panel C).

Only open-weight models were tested; generalization to frontier cloud models is unknown.

Six interface conditions were tested as single-turn prompts in the original seven-model subanalysis only; the thirteen added models were not evaluated under P1-P5. Designs that change the interaction rather than the instruction (autocomplete, turn-taking, pre-send confirmation) were not tested; the candidate-list figure bounds what such a step could recover, not a user performing it. The physician-validation panel (eMethods S6) covers only the main P0 run, so interface-condition results carry the same measurement uncertainty without a condition-specific human check or the CER-stratified correction computed for P0 (eMethods S8). Verbalized confidence is not comparable in scale across models, so the pooled calibration figure was descriptive, not a single transportable estimate.

Three chain-of-thought reasoning models ran on a reduced-replicate exposure grid (Methods) because the full grid was computationally intractable within budget, so their drift estimates carry wider confidence intervals (Results; eMethods S1, eTable 2); greater apparent reasoning capacity was not uniformly associated with lower drift among them.

Between-model ranking is descriptive checkpoint-as-served heterogeneity, not a controlled comparison of model size, reasoning capability, or architecture: checkpoints differed in quantization, serving configuration was unconfirmed for seven of thirteen expansion models, and inference-library version, chat-template, and context-window size were not recorded for any model (eTable 14).

## Conclusion

In this in-silico benchmark, language-model post-editing of simulated speller output produced fluent semantic substitutions whose rate rose with corruption and confidence did not reliably flag. Whether and how often this occurs in a patient-facing system will require prospective, human-in-the-loop study.

## Data and code availability

Analysis code, the frozen prompt bank, the frozen exposure file, and the result digests are available at https://github.com/BRIDGE-GenAI-Lab/BCI-Intent-drift-and archived as a public archive on the Open Science Framework. bigP3BCI is an open dataset available from PhysioNet; the Boston Children’s Hospital ALS Message Banking vocabulary is cited, not redistributed.

## Reporting

TRIPOD-LLM and the LLM-BCI reporting checklist from Gorenshtein et al, 2026.

## Data Availability

All data produced are available online at https://github.com/BRIDGE-GenAI-Lab/BCI-Intent-drift-

