## appendix for "Intent Drift in LLM-Assisted BCI Communication: An In-Silico Benchmark Under Simulated Decoder Corruption"

This appendix accompanies the main manuscript. It gives the full derivation of the P300 character-confusion corruption model, the corpus and matched-control construction, the outcome taxonomy and the automated evaluator ensemble, the physician validation panel, the complete statistical analysis, and the per-corpus, per-model, and calibration results that the main text summarizes. It reports numbers only and does not extend the clinical claims made in the main text. eTable 1 gives a dated timeline of the study's development.

### Contents

- eMethods. S1. Corpus sampling and Monte Carlo power. S2. P300 confusion-matrix corruption model. S3. Matched-control construction. S4. Outcome taxonomy, fluency gate, and critical-error detectors. S5. Multi-evaluator ensemble. S6. Physician validation panel. S7. Statistical analysis. S8. Interface-condition substudy. S9. Literature search.
- eTable 1. Analysis development timeline.
- eTable 2. Per-model drift rate and confidence calibration, current-generation panel.
- eTable 3. Drift rate by corpus and target character error rate, target-CER dose-response odds ratios, and target-CER two-way clustering and per-model slopes.
- eTable 4. Matched CRIT-versus-CTRL conditional logistic regression (rule-inclusive).
- eTable 5. Sensitivity of the drift estimate to tie-break rule and fluency threshold.
- eTable 6. Automated-ensemble-versus-physician-panel agreement.
- eTable 7. Interface-condition substudy.
- eTable 8. Message-deduplicated primary corruption slope, original seven-model realized-CER (Panel A) and current twenty-model target-CER (Panel B).
- eTable 9. Rule-free (detector-removed) matched CRIT-versus-CTRL conditional logistic regression.
- eTable 10. Drift by realized character error rate, and uniform-grid versus operating-CER-reweighted pooled drift.
- eTable 11. Drift-specific confidence calibration and high-confidence miss rate, all 20 models.
- eTable 12. Per-model realized-corruption slopes (20 models), two-way cluster-robust inference, and collinearity and mediation.
- eTable 13. Per-study decoder corruption, corruption-model allocation and structure, and held-out-participant matrix sensitivity.
- eTable 14. Model manifest for all 20 models: served tag, quantization, ollama config digest, immutable model-layer digest, and hardware/configuration confirmation.
- eTable 15. LLM-BCI reporting checklist crosswalk (Gorenshtein et al, ref 4).
- eFigure 1. Realized versus target character error rate.
- eFigure 2. Pooled 36-by-36 P300 character confusion matrix.
- eFigure 3. Per-model confidence-reliability small multiples.
- eFigure 4. Pooled confidence-outcome calibration (descriptive).
- Code and data availability.

### eMethods

#### S1. Corpus sampling and Monte Carlo power

Three message sets entered the exposure: the ALS message-banking vocabulary (AUTH), a message-critical challenge set (CRIT), and a matched control set (CTRL, constructed as described in S3). AUTH comprised the full Costello/Boston Children's Hospital collection, all 2,168 phrases across 40 categories, analyzed as a complete census rather than a sample drawn from it.

This design, 2,168 messages, five corruption levels, and 20 replicate corruptions per message-by-error-rate cell, was evaluated by a Monte Carlo power calculation for the primary drift-versus-corruption slope, anchored to the effect size observed in pilot data; it returned power of at least 0.99 to detect the anchored slope.

CRIT comprised 131 message-critical items, constructed and frozen before the main seven-model run began. Items spanned five categories in which a reconstruction error changes clinically consequential content: negation, numeral or dose, recipient, urgency, and actionable omission. AUTH and CRIT draw from different message populations and answer different questions, typical-case fidelity versus fidelity on disproportionately consequential content, so results are reported separately by corpus throughout rather than pooled into a single rate.

Three additional models (gpt-oss:20b, gpt-oss:120b, and deepseek-r1:32b), added to extend the panel from 16 to 20, produce substantially longer outputs than the rest of the panel because each interleaves an extended chain-of-thought reasoning trace before its final answer; serving them at the full corpus's replicate depth was not computationally tractable within the study's wall-clock budget. For these three models only, the exposure grid was reduced in replicate depth rather than in message or corruption-level coverage: every target CER level (0, 10, 20, 30, and 40 percent) is represented by an identical count of rows (8,100 each), and 2,299 of the corpus's 2,430 messages (94.6 percent) appear at least once, but the number of independent replicate corruptions drawn per message-by-target-CER cell was thinned from 20 to a deterministic subset of approximately 3 to 4 (selected by a fixed stride over the replicate index, not by discarding whole messages or whole corruption levels). The reduced grid totals 40,500 exposures and preserves the full corpus's AUTH-CRIT-CTRL composition to within 0.1 percentage points (89.2/5.4/5.4 percent reduced versus 89.2/5.4/5.4 percent full). Because coverage of messages, corruption levels, and corpus composition is preserved and only replicate depth is cut, the reduction is expected to widen the confidence intervals around these three models' drift estimates without introducing a systematic bias; per-model estimates for these three models carry this caveat throughout, and any panel-level pooled or weighted statistic accounts for the resulting unequal per-model N (eTable 2).

#### S2. P300 confusion-matrix corruption model

Decoder noise was parameterized by an empirical character-confusion matrix rather than by an assumed error distribution. The matrix was derived from bigP3BCI, an open dataset of P300-speller sessions. bigP3BCI as a whole includes both ALS and non-ALS control sub-studies, but derivation of the confusion matrix was restricted to the three constituent studies, labeled F, L, and N, that used the canonical 6-by-6 BCI2000 speller grid, a 36-symbol alphabet of the letters A through Z, the digits 1 through 9, and space; all 29 participants who contributed to those three studies were ALS-status, so the matrix was estimated from ALS participants only and contains no control participants (ALS status re-derived directly from each recording's patient-identification header; eTable 13). Each grid symbol maps to a row and a column index, and the linear symbol index was computed as (row minus 1) times 6 plus column, allowing the intended and selected targets of each recorded selection to be recovered from the stored grid codes.

Each selection's intended and selected target were reconstructed and pooled into a single 36-by-36, row-normalized matrix giving the probability of the selected character conditional on the intended character. The pooled matrix drew on 2,537 eligible selections from 29 subjects across 425 recording files; files that could not be parsed or reconstructed were skipped and counted. The empirical overall character error rate of the pooled matrix was 0.2136, with per-study values of 0.221 (study F, 10 participants, 1,067 selections), 0.161 (study L, 11 participants, 990 selections), and 0.306 (study N, 8 participants, 480 selections; eTable 13). The pooled matrix, rather than per-subject or per-study matrices, was used to corrupt the corpora: per-subject matrices were too sparse to support arbitrary text. A per-study drift sensitivity is reported by holding the AUTH drift-versus-realized-CER curve fixed and reweighting it by each study's own selection-count-weighted operating-CER distribution (eTable 13); a full per-study regeneration of the exposure panel from study-specific confusion matrices was not attempted given the added computational cost and is left as a bounded follow-up.

The confusion structure showed substitution mass concentrated within the same speller row or column as the intended symbol, the pattern produced by the row-and-column flash sequence of P300 spelling. This emergent structure is consistent with a matrix that reflects real P300 selection behavior rather than an arbitrary distribution; it does not establish the matrix as a criterion standard for any downstream label. eFigure 2 shows the pooled matrix.

Each message was corrupted at five target character error rates (CER): 0, 10, 20, 30, and 40 percent, with the 0 percent level a no-corruption reference. Messages were uppercased to match the speller alphabet, then corrupted by sampling substitutions from the pooled matrix; a fixed minority of the corruption budget was instead realized as single-character insertions or deletions. Characters outside the 36-cell grid passed through uncorrupted. Where a confusion row was empty, meaning a grid character was never observed as an intended target in the anchor data, a uniform substitution across the other grid characters was used as a fallback, so the intended error rate was still delivered.

Twenty independent replicate corruptions were drawn per message-by-target-rate cell. Every draw was seeded deterministically from a SHA-256 hash of the message identifier, the target CER, and the replicate index, rather than from a salted or session-dependent hash, so every realized corruption is exactly reproducible. Because the corruption draw is stochastic, the realized character error rate of each draw, not the target it aimed at, was used as the analytic exposure in every downstream model. The realized rate approximated its target at every level (for example, a target of 20 percent realized approximately 17.5 percent, and a target of 40 percent realized approximately 37.6 percent). eFigure 1 plots realized values against targets.

#### S3. Matched-control construction

Each of the 131 CRIT items was matched, one to one, to a non-critical control message (CTRL, n = 131) drawn from the remaining corpus. Matching used a greedy nearest-neighbor algorithm over five covariates: character length, word count, presence of a numeral, presence of a negation, and mean word frequency. Matching proceeded item by item, drawing the nearest unused candidate on the combined covariate distance for each CRIT item without replacement, and the standardized mean difference between the matched CRIT and CTRL sets was computed on all five covariates as the balance diagnostic.

Matched status made CTRL, not AUTH, the comparator for the message-critical contrast. With CTRL as comparator, any content-driven excess in CRIT could be evaluated after removing length, word count, numeral presence, and negation presence as confounders, isolating the contribution of message-critical status itself. CTRL was corrupted under the identical schedule as its paired CRIT item, at the same five target CER levels and the same 20 replicates, so the pairing survives into the exposure grid and supports the matched conditional-logistic comparison in S7.

#### S4. Outcome taxonomy, fluency gate, and critical-error detectors

Every reconstruction received one of three mutually exclusive labels, faithful, degraded, or drift, together with an independent fluency flag. An output was fluent if it read as ordinary, well-formed language, independent of whether it was correct. A faithful output was fluent and conveyed the intended message. A degraded output was disfluent or incomplete and asserted no different meaning from the intended message. A drift output was fluent but conveyed an intent different from the intended message, so it read as a plausible reconstruction while being wrong. The fluency gate was applied before the faithful-versus-degraded distinction could be made, so a fluent-but-wrong output was never conflated with a garbled, non-fluent one under the same label.

Meaning difference, the criterion separating drift from the other two labels, was adjudicated with a bidirectional natural-language-inference (NLI) contradiction check, a cosine-similarity threshold between the reconstruction and the intended message, and five rule-based detectors matched to the CRIT categories: a negation-flip detector, a numeral-or-dose-change detector, a recipient-change detector, an urgency-change detector, and an actionable-omission detector. The NLI check was retained because a negation flip can produce a high cosine similarity between output and intended message while remaining a logical contradiction, so similarity alone would miss one of the most consequential drift categories.

Message-critical status, membership in CRIT, is a fixed property of the input assigned before corruption; it is not an outcome label. A message-critical item could resolve as faithful, degraded, or drift like any other item, and its status entered the analysis only as a covariate, described in S7.

#### S5. Multi-evaluator ensemble

Outcome labels were produced by a multi-evaluator ensemble rather than a single fixed rule. Two natural-language-inference checkpoints (microsoft/deberta-large-mnli and roberta-large-mnli, applied in both directions) and two sentence-embedding checkpoints (all-mpnet-base-v2 and all-MiniLM-L6-v2, cosine threshold 0.75) formed four voting members, each combining its NLI and embedding signals with a shared fluency model (gpt2 perplexity rescaled to the unit interval, judged fluent at a threshold of 0.5) and five rule-based safety-relevant-subtype detectors into one of faithful, degraded, or drift. Meaning was always compared against the intended message. Disagreement among the four members was resolved by a tie-break rule fixed before the main seven-model run: a majority vote, with the deberta-mpnet member breaking a two-to-two tie. Full checkpoint identifiers, thresholds, the label-combination logic, and handling of empty or malformed outputs are in the released ensemble specification.

Mean pairwise inter-evaluator agreement across the ensemble was 0.979, with a tie rate of 3.9 percent requiring the tie-break rule; because the four members share related architectures and training data, this agreement reflects internal consistency, not independent validation. This ensemble produced the labels used throughout the primary analysis. The ensemble was evaluated, not validated: its output is a measured label subject to its own error rate, not a criterion standard, and its performance is being characterized against a blinded author-rater panel (S6) rather than assumed.

#### S6. Physician validation panel

Two physicians independently labeled a powered sample of 1,295 outputs: a stratified draw of 980 message-critical (CRIT) outputs, so that agreement on the category the study cares most about is measured directly, plus a stratified sample spanning the full CER grid and the full model panel, so that agreement is characterized across corruption levels and across models; across the full 1,295-item panel this placed 259 items at each CER level and 185 with each model. Physicians labeled blinded to model identity, corruption level, corpus, and the automated label, using the same four-level scale (faithful, degraded, drift, and message-critical drift). A third physician adjudicated the items on which the first two disagreed. The full message-critical census was 4,585 unique message-by-model-by-CER cells; rating every one was infeasible for a two-physician panel, so the panel used the powered draw targeting at least 1,000 critical items rather than a full census.

Two results were reported. First, class-specific agreement between the automated ensemble and the physician consensus (sensitivity, specificity, positive and negative predictive value, F1, and the full confusion structure, by outcome class; eTable 6). Second, a misclassification-corrected drift estimate that applies a Rogan-Gladen correction to the full-cohort automated rate using the panel-measured error rates of the ensemble (S7). Consensus was the shared label where the two physicians agreed and the third physician's label where they disagreed; every rated item, including the 156 that required adjudication, was carried into the automated-versus-consensus comparison, so the comparison denominator equals the full rated sample.

**Sixteen-model panel (primary measurement-validation analysis; adjudication complete). After the earlier seven-model panel was frozen and analyzed, a second two-physician panel rated a stratified extension spanning all 16 models, all 3 corpora, the full CER grid, and every automated outcome class (2,133 items), plus a dedicated zero-realized-CER sheet (148 items), for 2,281 blinded items in total (instrument and rating sheets archived in the study repository). Physician-versus-physician agreement across this full panel was Cohen kappa 0.759 (raw agreement 85.0%), consistent with the earlier seven-model panel's 0.79. A third physician (O.D.), who was not one of the two raters, then adjudicated the 343 items on which the raters disagreed, resolving all 2,281 items. Over the full resolved set (1,938 concordant plus 343 adjudicated), ensemble-versus-physician-consensus agreement was Cohen kappa 0.412 (moderate; on the 1,938 concordant items alone it was 0.434), lower than the earlier seven-model panel's 0.67 but reproducing the same class pattern: faithful validated well (F1 0.91), drift decently (F1 0.76), and degraded poorly (F1 0.52, the ensemble relabeling roughly half of physician-labeled degraded outputs as drift), the same weakness already reported for the seven-model panel above. With adjudication complete, the weighted misclassification (Rogan-Gladen) correction for this panel is available: corrected drift prevalence was 27.9% in AUTH (95% CI 22.1 to 33.0; from a naive automated rate of 31.3%), 33.4% in CRIT (21.7 to 43.3), 29.6% in CTRL (21.1 to 38.9), and 28.3% pooled (22.9 to 33.0). The same weighted correction was also computed stratified by target CER rather than by corpus, applying each level's own confusion matrix rather than the pooled one, using the identical stability guard (_stratum_confusion_is_stable, minimum 30 rated items with nonzero support for all three classes); every native CER level passed this bar, with 526, 453, 435, 435, and 432 rated items at 0%, 10%, 20%, 30%, and 40% target CER respectively (eTable 6, Panel F). This sixteen-model panel (eTable 6 Panel E) is the primary measurement-validation analysis, covering 16 of the study's full 20-model population (it predates the four models added last, eMethods S1; Discussion, Limitations); the earlier seven-model panel (eTable 6 Panels A-D) is reported as a further sensitivity analysis. Within the dedicated zero-CER sheet, two physicians independently confirmed a drift-type label on only 29.7% of the 148 items (rater 1, 37.8%; rater 2, 29.7%; both raters agreeing, 29.7%), and both independently called 57.4% (85 of 148) faithful, with physician-versus-physician kappa 0.752 (raw agreement 85.8%) on this subset; this two-rater result reinforces, with a number, the existing caveat that the automated zero-CER cause taxonomy is rule-based rather than physician-adjudicated. Class-specific and zero-CER figures for this panel are reported in eTable 6, Panel E.**

#### S7. Statistical analysis

The primary model was a mixed-effects logistic regression of the binary drift outcome (drift versus the other two outcomes) on realized corruption and message-level covariates, character length, error count, and flags for whether the realized corruption touched a negation token or a numeral, with crossed random intercepts for message and for model family, fit as a Bayesian cross-classified binomial mixed model using the statsmodels variational-Bayes implementation. At full scale (4,252,326 generations) that variational-Bayes fit did not converge. The estimate reported as primary throughout the manuscript is therefore a message-clustered, cluster-robust logistic regression on the same fixed-effect predictors, which converged but does not return message- or subject-level variance components.

The excess drift attributable to message-critical content was estimated separately, using the CRIT-CTRL pairing from S3. A matched-pair conditional logistic regression conditioned on the matched pair, target CER, replicate index, and model, and adjusted for the realised corruption each item actually received (realised CER, corrupted-character count, and corrupted numeral and negation indicators). Because a CRIT message and its CTRL match are different character strings, they do not receive identical realised corruption even at the same target CER and replicate index; the realised-corruption covariates adjust for that difference rather than assuming it away. Character length, word count, numeral presence, and negation presence were balanced by construction of the matched pair. The criticality contrast is estimated from the discordant pairs. Coefficients are reported in eTable 4.

Confidence calibration was summarized per model by the expected calibration error (ECE) and by the area under the receiver-operating-characteristic curve for verbalized confidence predicting a faithful output, denoted AUROC(confidence-to-faithful), each with a message-clustered bootstrap confidence interval. Because verbalized confidence is not comparable in scale across models that differ in size and training data, the cross-model summary used DerSimonian-Laird random-effects pooling rather than a fixed-effect estimate.

Because the safety argument turns on whether confidence flags drift specifically, and AUROC(confidence-to-faithful) tests only faithful against the pooled remainder, a drift-focused calibration companion was added per model (eTable 11): a drift-specific AUROC, in which a low-confidence score (1 minus stated confidence) predicts the drift label against all non-drift outputs (faithful and degraded pooled); the Brier score and Cox logistic recalibration intercept and slope of that same drift-risk score against the observed drift indicator; and the drift false-negative rate at confidence thresholds of 0.7 and 0.9, defined as the fraction of true drift outputs a "flag when confidence is below the threshold" rule would fail to flag. These metrics were computed within each model and never pooled across models on the raw confidence scale.

A misclassification-corrected drift estimate uses a multiclass Rogan-Gladen correction. It adjusts the full-cohort automated rate for the automated pipeline's own measured class-specific error rates rather than reporting the raw automated rate as though it carried no measurement error. A single pooled confusion matrix, estimated on the physician-rated validation panel (the pipeline is one labeler, so its error pattern is a property of the labeler), was applied to each corpus's naive label counts; the same pooled matrix drove both the point estimate and the message-clustered bootstrap interval. Estimating a separate matrix per corpus was rejected because per-corpus validation subsets can be small (as few as 70 items for CTRL in the earlier seven-model panel) and give unstable 3-by-3 inversions; the assumption that the confusion pattern is corpus-independent is a limitation. Applied to the primary sixteen-model panel (2,281 items), the correction lowered the detected drift rate in every corpus, pooled 31.4% to 28.3% and AUTH 31.3% to 27.9% (eTable 6, Panel E). Applied to the earlier seven-model sensitivity subset (1,295 items), the same correction instead raised the detected rate in every corpus (AUTH 30.9% to 36.4%), with wide, overlapping intervals (eTable 6, Panel D).

Sensitivity analyses varied two analyst choices in the labeling pipeline: the ensemble's tie-break rule and the fluency threshold used to separate degraded from faithful outputs. Results are reported in eTable 5. All corruption draws and model queries used deterministic seeds, and analysis code and the frozen exposure file are released alongside the manuscript. Because CER was manipulated experimentally rather than observed, its association with drift is reported as a controlled experimental estimate, not as an observational association.

#### S8. Interface-condition substudy

The six interface conditions are the six prompts of the bank frozen before the main seven-model run and fixed by SHA-256 hash (eTable 1). Forced reconstruction (P0) instructs the model to reconstruct the intended message without adding, removing, or changing meaning; minimal edit (P1) restricts it to correcting spelling; copy when uncertain (P2) instructs it to return the input unchanged unless a character is almost certainly an error; abstention permitted (P3) instructs it to return the sentinel "[ABSTAIN]" rather than guess when it cannot reconstruct with high confidence; candidate list (P4) requests one best reconstruction plus up to two alternatives; expansion (P5) requests the full fluent sentence the person most likely intended. The five non-P0 prompts were run as a full interface-condition substudy against all seven models.

The substudy subset was drawn from the main exposure grid: 300 AUTH messages, all 131 message-critical probes, and their 131 matched controls, at all five target corruption levels with 10 replicates each, for 28,100 exposures. Each was run under P1 through P5 against the same seven models, giving 983,500 generations. Forced reconstruction was not regenerated. Its rows were taken from the already-labeled main run at the identical exposures, so the reference arm uses byte-identical inputs rather than a fresh sample. Reuse is matched on the message-and-corpus pair rather than on message identifier alone, because the matched controls are drawn from the same message bank as AUTH and share identifiers with it, while the message-critical probes use distinct, disjoint identifiers; matching on identifier alone admits the AUTH twin of every reused control item and unbalances the reference arm. With pair matching the six arms are balanced at 196,700 generations each.

Abstention sits outside the outcome taxonomy of S4. A declined output was never passed to the labeler and never entered the faithful, degraded, or drift denominator, so no declined row is scored as a non-drift success by default. Because declining removes the hardest exposures from that denominator, every rate conditional on answering is reported alongside a throughput rate that counts declines (eTable 7, Panel A). Condition effects on drift were estimated by message-clustered cluster-robust logistic regression with forced reconstruction as the reference, fitted with Newton and an L-BFGS fallback. Both a condition-by-corruption interaction model and a main-effects model were fitted. The reported odds ratios are the main-effects terms, which average over the corruption grid; a condition term in the interaction model is instead the odds ratio at zero corruption, where drift is rare, and for copy when uncertain the two point in opposite directions. The corruption-resolved rates are reported directly in eTable 7, Panel B rather than as interaction coefficients. Benefit-harm transitions reuse the S5 transition definitions with condition added as a grouping key.

Candidate-list (P4) parsing runs in two stages. The raw reply is parsed as JSON, first as the whole string, then, if that fails, as the greedy brace-matched substring within it; if this yields a dict with no "candidates" key at all, that is read as a clean, genuine absence of alternates. Any other outcome (the reply is not valid JSON at either stage, or a "candidates" key is present but is not a list, or contains no usable non-empty-string entries) is treated as a malformed candidate list, not a genuine absence, and triggers a text-level fallback: a regex recovers a well-formed "candidates": [...] array directly from the raw text regardless of whether the surrounding JSON is broken, and extracts its quoted string elements. Only if that regex also finds nothing does the row fall back to having no candidates. The primary answer is parsed independently of this candidate-array logic, through the same "message"-field-or-whole-reply fallback every other condition uses, so a malformed candidate list never drops the row from the substudy or its faithful, degraded, or drift denominator; it only leaves that one row without recoverable alternates.

Each candidate is scored individually, not inferred from the primary answer. The primary answer (the "message" field) is labeled by the same evaluator ensemble as every other condition (eMethods S5): bidirectional natural-language-inference entailment and contradiction against the intended message, cosine similarity from two embedding models, and the fluency gate, combined by majority vote across the four evaluator combinations. Every alternate in the "candidates" array is separately paired with the same intended message and passed through that identical ensemble, so an alternate is labeled faithful, degraded, or drift on its own merits. The candidate-list recovery ceiling in eTable 7, Panel D, is built directly from these two label sets: a row's alternates are counted as recovering a faithful outcome when the primary answer is not itself labeled faithful but at least one separately scored alternate is, and the reported lift is the gap between that combined rate and the primary-answer-alone faithful rate. It is reported as a ceiling, not an achieved rate, because no user in this benchmark inspected the alternatives and selected among them.

Expansion (P5) output is compared against the same terse intended message using the identical, unmodified pipeline: the same two natural-language-inference models, the same two embedding models' cosine similarity, and the same fluency gate, with no length-based rescaling or other adjustment for P5's systematically longer output. A longer, elaborated reconstruction is therefore held to the same meaning-preservation and fluency criteria as a same-length one; expansion's higher drift and lower rescue in eTable 7 reflect that unmodified scoring, not a more lenient standard applied to its longer output.

The physician-validation panel (eMethods S6) was drawn entirely from the main run: every sampled item carries a message identifier and model from the seven- or sixteen-model main cohort, and the panel's key file carries no interface-condition or prompt identifier, because no interface-substudy row was ever eligible for sampling into it. Automated-versus-physician agreement, reported in eMethods S6 and eTable 6, was therefore never independently checked against P1-P5 outputs specifically. The interface-condition drift rates above, including the P4 candidate-recovery ceiling and the P5 expansion comparison, carry the same automated-pipeline measurement uncertainty as the main results, but without a condition-specific physician check and without the CER-stratified physician-consensus correction computed for the main P0 pipeline (eTable 6, Panel F).

#### S9. Literature search

The evidence-synthesis statement in the Research in context section of the main text (Evidence before this study) draws on a structured literature search that attempted five databases (PubMed, Scopus, IEEE Xplore, arXiv, and bioRxiv) and successfully queried three of them (PubMed, arXiv, and bioRxiv), each searched from database inception to July 19, 2026, with no restriction by publication language; Scopus and IEEE Xplore could not be queried, for the reason stated below. The search combined three term groups with Boolean AND: large language models; brain-computer interfaces and P300 spellers; and augmentative and alternative communication (AAC), the same term groups already named in the Research in context. This search also served as the basis for our related 2026 systematic review of language-model-integrated brain-computer interfaces, which named intent drift as a plausible but unmeasured failure mode.

The exact strings, adapted to each database's supported search syntax, were as follows.

- **PubMed:** ("large language model*"[tiab] OR "large language models"[tiab] OR LLM[tiab] OR LLMs[tiab] OR GPT*[tiab] OR "generative pretrained transformer*"[tiab] OR "foundation model*"[tiab]) AND ("brain-computer interface*"[tiab] OR "brain computer interface*"[tiab] OR BCI[tiab] OR BCIs[tiab] OR "P300 speller*"[tiab] OR "P300-based speller*"[tiab] OR "augmentative and alternative communication"[tiab] OR AAC[tiab])
- **Scopus:** TITLE-ABS-KEY("large language model*" OR "large language models" OR LLM OR LLMs OR GPT* OR "generative pretrained transformer*" OR "foundation model*") AND TITLE-ABS-KEY("brain-computer interface*" OR "brain computer interface*" OR BCI OR BCIs OR "P300 speller*" OR "P300-based speller*" OR "augmentative and alternative communication" OR AAC)
- **IEEE Xplore:** ("Full Text & Metadata":"large language model*" OR "Full Text & Metadata":"large language models" OR "Full Text & Metadata":LLM OR "Full Text & Metadata":LLMs OR "Full Text & Metadata":GPT) AND ("Full Text & Metadata":"brain-computer interface*" OR "Full Text & Metadata":"brain computer interface*" OR "Full Text & Metadata":BCI OR "Full Text & Metadata":"P300 speller*" OR "Full Text & Metadata":"augmentative and alternative communication")
- **arXiv:** (large language model OR large language models OR LLM OR LLMs OR GPT) AND (brain-computer interface OR brain computer interface OR BCI OR P300 speller OR augmentative and alternative communication OR AAC), entered as a free-text abstract search; arXiv's search interface has no field-restricted Boolean syntax equivalent to PubMed or Scopus.
- **bioRxiv:** the same term combination as arXiv, entered as a free-text abstract search; bioRxiv's own search interface has no field-restricted Boolean equivalent and, in this environment, could not be reached directly (see the count paragraph below), so the equivalent query was run against Europe PMC's bioRxiv-restricted index instead.

Record counts were obtained by actually executing the exact strings above against each database's live API or interface, with an explicit upper date bound applied so the returned counts match the July 19, 2026 window stated above, rather than being estimated or reconstructed. Three of the five named databases were reachable this way: PubMed, 52 records (NCBI E-utilities esearch); arXiv, 80 records (arXiv API); and bioRxiv, 5 records. bioRxiv's own search interface returned a bot-protection challenge to every automated access attempt and could not be queried directly; the bioRxiv figure was instead obtained from Europe PMC's index of bioRxiv preprints, a real, independently queryable index of bioRxiv content, not bioRxiv's own search box, and should be read with that caveat. Scopus and IEEE Xplore could not be queried at all: both require an institutional subscription or API credential not available in this environment, and neither is reported as a number here, invented or otherwise. The identified total across the three reachable databases was 137 records, covering three of the five named databases, not all five.

Of these 137 records, 128 remained after removing 9 confirmed duplicates, found by exact-title matching plus, on the subset of visibly similar remaining titles, exact author-list verification of three further preprint-to-published-paper pairs. All 128 deduplicated records were screened by title and abstract. Sixty were judged topically relevant, meaning their title and abstract describe a large language model or foundation model used within, or evaluated against, a brain-computer interface, P300 speller, or augmentative-and-alternative-communication pipeline; this is a title-and-abstract relevance judgment, reconstructed after the fact from the retrieved title lists rather than logged at the time of the original search, and no full text was retrieved or read for any record. Six of these 60 were both cited in the evidence-before-this-study passage of the Research in context section and confirmed present among the 128 deduplicated records by exact DOI or arXiv identifier, informing the evidence synthesis. Three further references cited in that same passage were checked and confirmed absent from the retrieved set, most likely because their titles and abstracts use terminology, for example "EEG-to-text," outside this search's literal term list; this is an honest limit on the search's sensitivity, not a gap in the counting exercise. Full per-database query provenance and the underlying record lists are archived alongside the analysis code.

### eTables

#### eTable 1. Analysis development timeline

A dated timeline of the study's development, from the initial pilot through the final 20-model analysis. Dates are established from the project's version-control history.

| Date | Development |
| --- | --- |
| 2026-07-19 | Endpoints, outcome taxonomy, model list, and CER grid established, following a 4-model, 5,020-generation baseline pilot; target CER set as the primary exposure alongside confidence calibration (AUROC, expected calibration error); replicate design set at 20 draws per message-by-CER cell; matched CRIT-CTRL controls and a fluency gate added to the outcome taxonomy. |
| 2026-07-20 | Main seven-model run generated, labeled, and analyzed (7 models x 76,200 exposures = 533,400 generations). Misclassification (Rogan-Gladen) correction and zero-corruption drift audit added. |
| 2026-07-23 to 2026-07-26 | Interface-condition substudy run as a full six-arm design on a 562-message subset against all seven models. |
| 2026-08-02 | Model panel expanded from 7 to 16 models (3,888,000 generations). Two-way (message-by-model) clustering, 16-model calibration, 16-model matched CRIT-CTRL excess, corruption-profile analysis, and the stratified 16-model physician-validation panel added; realized CER reported as a secondary dose-response alongside the primary target-CER analysis. |
| 2026-08-11 | Model panel expanded from 16 to 20 models by adding granite3.3:8b and three chain-of-thought reasoning models (gpt-oss:20b, gpt-oss:120b, deepseek-r1:32b), the latter three evaluated on a reduced-replicate 40,500-exposure grid rather than the full 243,000-exposure grid because of their added inference cost (eMethods S1). Twenty-model calibration, matched-CRIT-CTRL excess, drift-specific calibration, target-CER dose-response, two-way clustering, and corruption-profile analyses re-run on the expanded panel (4,252,326 generations); the physician-validation panel (eMethods S6) remains on the earlier 16-model panel (Discussion, Limitations). |

The primary mixed-effects model for the drift-versus-CER dose-response was specified as a Bayesian cross-classified binomial mixed model; that engine's variational-Bayes fit did not converge at full scale, so the estimate reported as primary is a message-clustered, cluster-robust logistic regression on the same predictors (eMethods S7).

#### eTable 2. Per-model drift rate and confidence calibration, current-generation panel

Drift rate in AUTH and per-model calibration for the full 20-model panel, ordered from lowest to highest drift. The meta-analytic row pools calibration across all 20 models by DerSimonian-Laird random-effects pooling; 15,814 outputs with missing verbalized confidence were dropped from the calibration calculations. That missingness was not at random with respect to drift: among the 15,814 dropped outputs the drift rate was 67.9%, against a 31.6% drift rate across the full panel (roughly a two-fold enrichment), so confidence was least available exactly where a drift-detection gate would need it most; this enrichment is smaller than the three-fold pattern seen in the earlier 16-model panel because the four added models do not share the earlier panel's missingness-drift association (gpt-oss:20b in particular shows the opposite pattern, its missing-confidence rows almost never drift). The full per-model missingness breakdown is in eTable 11's missingness audit. The interface-condition substudy elsewhere in this supplement remains limited to the original seven-model panel. The primary physician-panel validation remains on the earlier sixteen-model panel (eMethods S6; eTable 6, Panel E) and does not cover the four models added last (granite3.3:8b, gpt-oss:20b, gpt-oss:120b, deepseek-r1:32b); the original seven-model panel (eTable 6, Panels A-D) is reported as a further sensitivity analysis.

| Model | Drift rate, AUTH (%) | ECE | AUROC (confidence-to-faithful) |
| --- | --- | --- | --- |
| GPT-OSS-120B | 20.8 | 0.120 | 0.899 |
| GPT-OSS-20B | 22.0 | 0.193 | 0.882 |
| Gemma-4-31B | 23.1 | 0.173 | 0.896 |
| Qwen-2.5-7B | 23.6 | 0.417 | 0.827 |
| Gemma-4-E4B | 24.1 | 0.325 | 0.876 |
| Gemma-3-27B | 27.1 | 0.329 | 0.741 |
| Llama-4-Scout | 27.3 | 0.271 | 0.766 |
| Qwen-3.6-27B | 28.0 | 0.265 | 0.853 |
| Command-R | 28.2 | 0.346 | 0.742 |
| Gemma-4-12B | 28.3 | 0.304 | 0.877 |
| Qwen-3.5-27B | 28.7 | 0.220 | 0.884 |
| Qwen-3.6-35B-A3B | 29.0 | 0.280 | 0.870 |
| Gemma-2-9B | 31.7 | 0.346 | 0.785 |
| Mistral-Small-24B | 32.3 | 0.323 | 0.815 |
| DeepSeek-R1-32B | 35.1 | 0.340 | 0.841 |
| Phi-4-14B | 36.4 | 0.351 | 0.873 |
| Granite-3.3-8B | 37.5 | 0.397 | 0.797 |
| Llama-3.1-8B | 40.1 | 0.400 | 0.834 |
| Phi-4-mini | 43.3 | 0.502 | 0.734 |
| Phi-3-mini | 49.3 | 0.578 | 0.710 |
| Meta-analytic (random-effects), all 20 models | N/A | 0.32 (95% CI, 0.27 to 0.37) | 0.83 (95% CI, 0.80 to 0.85) |

Four models (granite3.3:8b, gpt-oss:20b, gpt-oss:120b, deepseek-r1:32b) complete the panel at 20; the three reasoning models among them (gpt-oss:20b, gpt-oss:120b, deepseek-r1:32b) are evaluated on a reduced-replicate 40,500-exposure grid rather than the full 243,000-exposure grid used for the other 17 models, because of their added inference cost from chain-of-thought reasoning; the reduced grid preserves full coverage of every message, corruption level, and corpus category (eMethods S1) and its estimates for these three models carry correspondingly wider confidence intervals. Notably, the two gpt-oss checkpoints have the lowest AUTH drift rate and lowest ECE in the entire panel despite (or because of) their reasoning traces, while deepseek-r1:32b, also a reasoning model, sits above the panel median on both, so chain-of-thought reasoning is not by itself associated with lower drift or better calibration.

**Generation-weighted versus equal-model-weighted pooling. Every pooled drift rate reported elsewhere in this manuscript, including the headline 31.5%/33.7%/31.5% AUTH/CRIT/CTRL rates above, is generation-weighted: each model's contribution to the pooled denominator is proportional to how many generations it produced (243,000 for the 17 full-grid models, 40,500 or 40,326 for the 3 reduced-replicate reasoning models), so a full-grid model implicitly outweighs a reduced-replicate model roughly six to one. As a sensitivity check on this panel-composition choice, re-pooling with each of the 20 models given one equal vote regardless of its generation count (the unweighted mean of the per-model AUTH rates in the table above) gives 30.8% in AUTH, 32.9% in CRIT, and 30.6% in CTRL, a small downward shift from the generation-weighted rates. The shift is downward because two of the three reduced-replicate models, gpt-oss:20b and gpt-oss:120b, are among the panel's lowest-drift checkpoints; giving them one vote each instead of a vote proportional to their smaller generation count up-weights them relative to generation-weighted pooling. Neither weighting is presented as more correct than the other; both are reported so a reader can judge how much the panel's own composition (17 full-grid models plus 3 reduced-replicate reasoning models) could move the headline number.**

#### eTable 3. Drift rate by corpus and target character error rate, the target-CER dose-response odds ratios, and target-CER cluster-robust and per-model slopes

Panel A gives the detected drift rate at each target CER, by corpus; this is the primary dose-response (eMethods S7). Panel B gives the overall outcome shares by corpus, pooled across all target CER levels. Panel C gives the two model-based summaries of the Panel A dose-response, both message-clustered and fit on the full 20-model, 4,252,326-generation panel: a binary (linear-in-target) logistic regression of drift on target CER, and a categorical logistic regression against the 0% reference. Panel D documents the target-CER uncertainty analyses cited in the main text: the pooled slope under message-only versus message-by-model two-way clustering, all 20 per-model target-CER slopes, their median and range, and the small-cluster caveat.

Panel A. Detected drift rate (%) by corpus and target CER.

| Target CER | AUTH | CRIT | CTRL |
| --- | --- | --- | --- |
| 0% | 2.2 | 3.0 | 1.7 |
| 10% | 14.4 | 17.2 | 15.5 |
| 20% | 31.9 | 35.5 | 32.4 |
| 30% | 48.6 | 51.4 | 48.1 |
| 40% | 60.3 | 61.3 | 59.7 |

Panel B. Overall outcome shares by corpus (%).

| Corpus | Faithful | Degraded | Drift |
| --- | --- | --- | --- |
| AUTH | 55.6 | 12.9 | 31.5 |
| CRIT | 50.6 | 15.7 | 33.7 |
| CTRL | 53.1 | 15.4 | 31.5 |

Panel C. Target-CER dose-response odds ratios, primary analysis (all corpora pooled, message-clustered, n = 4,252,326; 2,299 message clusters).

The binary (linear-in-target) model gave a log-odds slope of 8.311 (95% CI, 8.218 to 8.404) per unit target CER, equivalent to an odds ratio of 2.30 per 10-percentage-point increase in target CER.

| Target CER (vs 0% reference) | Odds ratio | 95% CI | P value |
| --- | --- | --- | --- |
| 10% | 7.71 | 7.00 to 8.48 | <.001 |
| 20% | 21.29 | 19.22 to 23.58 | <.001 |
| 30% | 42.71 | 38.46 to 47.43 | <.001 |
| 40% | 68.27 | 61.35 to 75.97 | <.001 |

The binary model used statsmodels.api.Logit (drift on target CER, cluster-robust by message); the categorical model used statsmodels.formula.api.logit with target CER entered as a categorical predictor against the 0% reference, also message-clustered.

Panel D. Target-CER dose-response under message-by-model two-way clustering, and per-model slopes (n = 4,252,326; 2,299 message clusters; 20 model clusters).

The pooled binary target-CER slope is unchanged (log-odds 8.311; odds ratio 2.30 per 10 percentage points). The standard error and 95% CI are reported below under message-only clustering and under Cameron-Gelbach-Miller message-by-model two-way clustering; the two-way interval is materially wider because model identity is a repeated cluster.

| Clustering | SE (log-odds slope) | Odds ratio per 10pp | 95% CI |
| --- | --- | --- | --- |
| Message-only | 0.048 | 2.30 | 2.27 to 2.32 |
| Message-by-model (two-way) | 0.332 | 2.30 | 2.15 to 2.45 |

Per-model target-CER slopes, each model fit separately (log-odds per unit target CER and the corresponding odds ratio per 10 percentage points), ordered from shallowest to steepest:

| Model | Slope (log-odds) | OR per 10pp |
| --- | --- | --- |
| qwen2.5:7b | 5.14 | 1.67 |
| deepseek-r1:32b | 5.43 | 1.72 |
| gpt-oss:20b | 6.34 | 1.89 |
| llama3.1:8b | 7.47 | 2.11 |
| gemma4:e4b | 7.57 | 2.13 |
| command-r | 7.97 | 2.22 |
| phi3:mini | 7.97 | 2.22 |
| gpt-oss:120b | 8.07 | 2.24 |
| llama4:17b-scout | 8.21 | 2.27 |
| granite3.3:8b | 8.41 | 2.32 |
| phi4:mini | 8.48 | 2.34 |
| qwen3.6:35b-a3b | 9.23 | 2.52 |
| gemma3:27b | 9.34 | 2.54 |
| mistral-small:24b | 9.45 | 2.57 |
| gemma4:12b | 9.61 | 2.61 |
| gemma2:9b | 9.71 | 2.64 |
| phi4:14b | 9.92 | 2.70 |
| qwen3.5:27b-q4 | 9.95 | 2.71 |
| qwen3.6:27b | 10.03 | 2.73 |
| gemma4:31b | 10.55 | 2.87 |

The median per-model slope was 8.45 (odds ratio 2.32 per 10 percentage points), range 5.14 to 10.55 (odds ratios 1.67 to 2.87). The two lowest-slope models are both reduced-replicate-grid reasoning models (deepseek-r1:32b, gpt-oss:20b), pulling the median down from the prior 16-model panel's 9.28; the third reduced-replicate model, gpt-oss:120b, is mid-pack (8.07), so this is not a uniform effect of the reduced grid across all three heavy models. Model-cluster inference is unstable with only 20 clusters, still below the 30-to-50-cluster rule of thumb: the two-way clustered interval is reported for honesty and correctly widens relative to the message-only interval, but with 20 clusters it should be read as approximate rather than a precise bound, and the per-model slopes with their median and range are the more informative view of model-level heterogeneity.

#### eTable 4. Matched CRIT-versus-CTRL conditional logistic regression (rule-inclusive)

Panel A. Original seven-model matched-pair conditional logistic regression.

Coefficients from the original seven-model matched-pair conditional logistic regression described in S7, conditioned on the matched pair, target CER, replicate index, and model, with adjustment for realised-corruption covariates (55,624 matched observations; 27,812 discordant pairs; model converged).

| Term | Coefficient (log-odds) | 95% CI | P value |
| --- | --- | --- | --- |
| Message-critical status | 0.144 | 0.118 to 0.171 | <.001 |
| Realized character error rate | 4.743 | 4.317 to 5.169 | <.001 |
| Corrupted-character count | 0.138 | 0.117 to 0.159 | <.001 |
| Corrupted numeral flag | 0.679 | 0.116 to 1.242 | .02 |
| Corrupted negation flag | 0.041 | -0.114 to 0.196 | .60 |
| Character length | 0.001 | -0.017 to 0.019 | .91 |

Exponentiating the message-critical coefficient gives an odds ratio of 1.16, matching the value reported in the main Results.

Panel B. Twenty-model pooled and per-model rule-inclusive matched odds ratio (the five critical-substitution detectors included in the drift label, matching Panel A's construction; the rule-free companion, with those detectors removed from the label, is in eTable 9). Odds ratios are from the same matched-pair conditional-logistic pipeline as Panel A, refit separately per model on that model's own matched pairs; the pooled row conditions on model as an additional stratum alongside the matched pair, target CER, and replicate index.

Pooled (all 20 models): OR 1.147 (95% CI, 1.129 to 1.166), 70,204 matched pairs.

| Model | Rule-inclusive OR | 95% CI | Matched pairs |
| --- | --- | --- | --- |
| Phi-3-mini | 1.01 | 0.96 to 1.07 | 4,827 |
| Gemma-4-E4B | 1.07 | 1.01 to 1.15 | 3,916 |
| Command-R | 1.08 | 1.01 to 1.15 | 4,218 |
| Qwen-2.5-7B | 1.09 | 1.03 to 1.16 | 4,093 |
| Gemma-2-9B | 1.11 | 1.04 to 1.19 | 4,033 |
| Qwen-3.5-27B | 1.12 | 1.04 to 1.20 | 3,882 |
| Gemma-3-27B | 1.14 | 1.06 to 1.22 | 3,791 |
| Phi-4-mini | 1.15 | 1.08 to 1.22 | 4,361 |
| Gemma-4-31B | 1.15 | 1.06 to 1.25 | 3,451 |
| Granite-3.3-8B | 1.16 | 1.08 to 1.24 | 4,272 |
| Qwen-3.6-27B | 1.16 | 1.08 to 1.25 | 3,790 |
| Phi-4-14B | 1.16 | 1.08 to 1.25 | 4,012 |
| Qwen-3.6-35B-A3B | 1.21 | 1.13 to 1.30 | 3,920 |
| Mistral-Small-24B | 1.21 | 1.13 to 1.30 | 4,194 |
| Llama-4-Scout | 1.25 | 1.17 to 1.34 | 4,033 |
| Gemma-4-12B | 1.25 | 1.17 to 1.35 | 3,996 |
| Llama-3.1-8B | 1.28 | 1.20 to 1.36 | 4,729 |

As in eTable 9's rule-free companion table, the three reduced-replicate-grid reasoning models (deepseek-r1:32b, gpt-oss:20b, gpt-oss:120b) do not appear here: their per-model conditional logistic regression failed to converge for the same reason (too little within-pair outcome variance once each model's matched pairs are limited to its own reduced-replicate grid; eTable 9, Panel B). The pooled 20-model estimate above still includes their matched pairs.

Panel C. Matched-pair covariate balance and match quality (the CRIT-CTRL matched-control construction, eMethods S3; shared by eTable 4 and eTable 9, since both reuse the identical 131 CRIT-CTRL pairs). Standardized mean difference (SMD) before and after matching, five covariates.

| Covariate | SMD before matching | SMD after matching |
| --- | --- | --- |
| Character length | 0.052 | 0.004 |
| Word count | 0.073 | 0.009 |
| Has numeral | 0.124 | 0.124 |
| Has negation | -0.254 | 0.000 |
| Mean word frequency | -0.570 | -0.025 |

All five covariates fall below the conventional 0.15 SMD threshold after matching. Has-numeral is unchanged from its pre-match value because it is a matching stratum (an exact-category variable), not a continuously balanced covariate. Match distance across the 131 pairs: median 0.099, mean 0.491, range 0.000 to 41.015 (the single largest distance is one atypical pair; the median and the after-match SMDs indicate the matching is close for the large majority of pairs). We attempted a single-extreme-match sensitivity analysis, refitting eTable 4 Panel A's and eTable 9 Panel A's identical seven-model matched design with this one pair (probe_0129, matched to costello_1875, standardized distance 41.01) excluded; dropping it left the conditional-logistic design numerically singular across five different optimizers, so no with-versus-without odds ratio could be reported for this exclusion under the same model specification (Discussion, Limitations).

#### eTable 5. Sensitivity of the original seven-model drift estimate to tie-break rule and fluency threshold

Panel A. Tie-break rule (all other settings at their primary value).

| Tie-break rule | Detected drift rate (%) |
| --- | --- |
| Primary rule | 31.0 |
| Drift-averse rule | 27.6 |
| Severity rule | 31.6 |

Panel B. Fluency threshold τ, separating degraded from faithful outputs (all other settings at their primary value).

| τ | Detected drift rate (%) |
| --- | --- |
| 0.3 | 34.7 |
| 0.4 | 33.2 |
| 0.5 (primary) | 31.0 |
| 0.6 | 28.3 |

#### eTable 6. Automated-ensemble-versus-physician-panel agreement

Panels A-D report an earlier seven-model validation subset (1,295 blinded items rated by two physicians, with a third adjudicating disagreements; eMethods S6), reported here as a sensitivity analysis. Kappa is Cohen kappa. Class-specific metrics compare the automated ensemble against the physician consensus label. Panel E reports the primary measurement-validation analysis, a later, complete sixteen-model panel (two raters plus adjudicator) covering 16 of the study's full 20-model population; it predates the four models added last to reach 20 (eMethods S1).

**Panel A. Physician-versus-physician and ensemble-versus-consensus agreement**

| Comparison | Raw agreement | Cohen kappa |
| --- | --- | --- |
| Physician versus physician, three-class label | 88.0% | 0.79 |
| Physician versus physician, faithful | -- | 0.93 |
| Physician versus physician, drift | -- | 0.72 |
| Physician versus physician, degraded | -- | 0.64 |
| Physician versus physician, message-critical subdistinction (both-drift items) | -- | 0.13 |
| Automated ensemble versus physician consensus, three-class label | 81.5% | 0.67 |

**Panel B. Automated ensemble versus physician consensus, class-specific agreement**

| Class | Sensitivity | Specificity | PPV | NPV | F1 | Support |
| --- | --- | --- | --- | --- | --- | --- |
| Faithful | 0.91 | 0.91 | 0.93 | 0.88 | 0.92 | 751 |
| Drift | 0.76 | 0.90 | 0.76 | 0.90 | 0.76 | 380 |
| Degraded | 0.49 | 0.91 | 0.45 | 0.93 | 0.47 | 164 |

**Panel C. Confusion matrix (physician-consensus rows, automated-ensemble columns), counts**

| Physician consensus | Auto faithful | Auto degraded | Auto drift |
| --- | --- | --- | --- |
| Faithful | 685 | 47 | 19 |
| Degraded | 12 | 81 | 71 |
| Drift | 37 | 54 | 289 |

**Panel D. Misclassification-corrected (Rogan-Gladen) drift versus detected automated rate**

| Corpus | Detected drift | Corrected drift | 95% CI |
| --- | --- | --- | --- |
| AUTH | 30.9% | 36.4% | 24.5-47.1 |
| CRIT | 33.4% | 36.2% | 20.6-50.7 |
| CTRL | 31.2% | 32.9% | 18.4-47.3 |

Correction raised rather than lowered the estimate in every corpus in this seven-model sensitivity subset, though with substantial uncertainty, so we do not claim these detected rates are definitively conservative; the correction assumes a corpus-invariant classification error. The primary sixteen-model panel (Panel E, below) shows the opposite direction, the correction lowering detected drift in every corpus. Intervals are message-clustered bootstrap (1,000 resamples); every point estimate lies within its interval, with no simplex clipping and no identity-row fallback.

**Panel E. Sixteen-model panel (two raters plus adjudicator; complete) -- primary measurement-validation analysis**

A sampling-weighted, blinded panel spanning all 16 models, all 3 corpora, the full CER grid, and every automated outcome class (eMethods S6). This sixteen-model panel is the primary measurement-validation analysis, covering 16 of the study's full 20-model population; it predates the four models added last to reach 20 (eMethods S1). The earlier seven-model panel (Panels A-D) is reported as a further sensitivity analysis. A third physician (O.D.), not one of the two raters, adjudicated the 343 disagreements, resolving all 2,281 items; the weighted Rogan-Gladen correction for this sixteen-model panel is shown below and lowers, rather than raises, the detected rate in every corpus.

| Comparison | Raw agreement | Cohen kappa |
| --- | --- | --- |
| Physician versus physician, three-class label (2,281 items) | 85.0% | 0.759 |
| Ensemble versus physician consensus, three-class label (all 2,281 resolved items; 1,938 concordant plus 343 adjudicated) | 60.7% | 0.412 |

On the 1,938 concordant items alone, ensemble-versus-consensus agreement was Cohen kappa 0.434 (raw 62.6%).

| Class | F1 (ensemble versus consensus, 2,281 resolved items) |
| --- | --- |
| Faithful | 0.91 |
| Drift | 0.76 |
| Degraded | 0.52 |

| Corpus | Naive automated drift | Rogan-Gladen corrected drift (95% CI) |
| --- | --- | --- |
| AUTH | 31.3% | 27.9% (22.1 to 33.0) |
| CRIT | 33.5% | 33.4% (21.7 to 43.3) |
| CTRL | 31.3% | 29.6% (21.1 to 38.9) |
| Pooled | 31.4% | 28.3% (22.9 to 33.0) |

The class pattern reproduces Panel B above: faithful and drift validate well, degraded poorly, with the ensemble again relabeling roughly half of physician-labeled degraded outputs as drift. Unlike the earlier seven-model sensitivity subset, where the same correction raised the AUTH estimate (30.9% to 36.4%; Panel D), this primary sixteen-model panel's correction lowers the detected rate in every corpus; the corrected rate nonetheless remains substantial.

Within the 148-item dedicated zero-realized-CER sheet, rated by both physicians:

| Zero-CER subset (148 items, two raters) | Value |
| --- | --- |
| Physician versus physician, Cohen kappa | 0.752 (raw agreement 85.8%) |
| Both raters confirmed a drift-type label | 29.7% (44/148) |
| Either rater confirmed a drift-type label | 37.8% (56/148) |
| Both raters called the item faithful | 57.4% (85/148) |

Both physicians independently rejected the automated drift label on most of these zero-realized-CER items, calling a majority of them faithful instead; this quantifies the existing caveat that the automated zero-CER cause taxonomy is rule-based rather than physician-adjudicated..

**Panel F. CER-stratified weighted misclassification correction**

The corpus-level correction in Panel E above pools the physician-measured confusion matrix across all five target-CER levels. To check whether the primary target-CER dose-response itself, not only the corpus-pooled correction, holds up against physician-consensus labels, the same weighted Rogan-Gladen correction was recomputed separately within each of the five target-CER strata, using each stratum's own confusion matrix rather than the pooled one (eMethods S6). Panel F's population is the 16-model panel pooled across all three corpora (AUTH, CRIT, and CTRL together), not AUTH alone; this is a different population from the AUTH-only, 20-model headline dose-response table in Results, which is why Panel F's naive rate at each target-CER level (this table's second column) differs slightly from that headline table's AUTH-only rate at the same level (for example 1.9% versus 2.2% at 0% target CER).

| Target CER | Naive automated rate | Corrected rate | 95% CI | Validation n |
| --- | --- | --- | --- | --- |
| 0% | 1.9% | 0.0% | 0.0-96.8 | 526 |
| 10% | 14.4% | 12.2% | 3.0-21.1 | 453 |
| 20% | 31.8% | 36.2% | 26.8-45.8 | 435 |
| 30% | 48.5% | 43.4% | 29.0-53.4 | 435 |
| 40% | 60.3% | 49.3% | 31.3-62.9 | 432 |

The corrected rate rose monotonically with target CER, reproducing the primary automated dose-response's direction at every level. The 0% level's 95% CI spans nearly the full probability range: with a near-zero true prevalence at that level, the Rogan-Gladen point estimate was clipped to the boundary of the probability simplex, so that stratum's correction is directionally consistent but not precisely estimated; the other four strata gave informative intervals. Every stratum met the same stability guard used for the corpus-stratified correction above (minimum 30 rated items with nonzero support for all three classes; eMethods S6).

#### eTable 7. Interface-condition substudy

Six interface conditions on a common subset of 562 messages (300 AUTH, 131 message-critical, 131 matched controls) at five target corruption levels, 10 replicates, and seven models: 196,700 generations per condition and 1,180,200 in total (eMethods S8). Forced reconstruction is the reference condition and was reused from the main run at identical exposures rather than regenerated. Declined outputs are counted separately and never enter the faithful, degraded, or drift denominator; the two "per 100 attempted" columns put declines back in the denominator, so they are comparable across conditions. Odds ratios are from a message-clustered cluster-robust logistic regression averaged over the corruption grid (562 clusters; 1,131,003 scored outputs after excluding 49,197 declines); every one's 95% CI in Panel A excludes 1 (p<0.001 throughout).

**Panel A. Outcome rates and adjusted drift odds by interface condition**

| Interface condition | Declined | Drift (of answered) | Faithful (of answered) | Drift per 100 attempted | Faithful per 100 attempted | Drift odds ratio vs forced (95% CI) |
| --- | --- | --- | --- | --- | --- | --- |
| Forced reconstruction (P0, reference) | 0.0% | 31.5% | 56.7% | 31.5 | 56.7 | 1 [reference] |
| Minimal edit (P1) | 0.0% | 28.6% | 55.9% | 28.6 | 55.9 | 0.84 (0.83-0.85) |
| Copy when uncertain (P2) | 0.0% | 26.9% | 54.6% | 26.9 | 54.6 | 0.75 (0.74-0.77) |
| Abstention permitted (P3) | 25.0% | 24.0% | 66.6% | 18.0 | 49.9 | 0.85 (0.83-0.86) |
| Candidate list (P4) | 0.0% | 33.6% | 55.0% | 33.6 | 55.0 | 1.13 (1.11-1.15) |
| Expansion (P5) | 0.0% | 35.3% | 55.5% | 35.3 | 55.5 | 1.25 (1.20-1.31) |

**Panel B. Drift rate (of answered outputs) by target character error rate**

| Interface condition | 0% | 10% | 20% | 30% | 40% |
| --- | --- | --- | --- | --- | --- |
| Forced reconstruction (P0, reference) | 1.2% | 13.1% | 31.4% | 48.9% | 62.2% |
| Minimal edit (P1) | 0.7% | 12.5% | 30.2% | 45.0% | 54.5% |
| Copy when uncertain (P2) | 1.1% | 13.1% | 29.4% | 42.0% | 48.6% |
| Abstention permitted (P3) | 1.3% | 11.9% | 28.3% | 44.3% | 57.5% |
| Candidate list (P4) | 4.7% | 15.5% | 32.6% | 50.4% | 64.8% |
| Expansion (P5) | 5.9% | 15.9% | 33.7% | 52.3% | 68.9% |

The decline rate under permitted abstention rose with corruption, from 5.6% at no target corruption to 10.1%, 20.8%, 36.4%, and 52.2% at 10, 20, 30, and 40 percent, so its detected-drift rates in this panel are conditional on a progressively more selected set of answered exposures.

**Panel C. Benefit-harm transitions against the raw decoded stream, pooled**

| Interface condition | Rescue | Silent failure | Model-induced harm | Faithful gained per fluent error |
| --- | --- | --- | --- | --- |
| Forced reconstruction (P0, reference) | 28.2% | 30.0% | 0.6% | 0.92 |
| Minimal edit (P1) | 27.3% | 27.2% | 0.4% | 0.99 |
| Copy when uncertain (P2) | 26.0% | 25.3% | 0.5% | 1.01 |
| Abstention permitted (P3) | 31.1% | 22.4% | 0.7% | 1.35 |
| Candidate list (P4) | 28.2% | 31.1% | 1.4% | 0.87 |
| Expansion (P5) | 28.5% | 32.6% | 1.7% | 0.83 |

Transition categories follow eMethods S5. A declined output has no reconstruction to compare against the raw decode, so declines are excluded from this panel; the abstention row is therefore computed on its answered exposures only.

**Panel D. Candidate-list recovery ceiling**

| Target CER | Primary answer faithful | Any candidate faithful | Recovery lift |
| --- | --- | --- | --- |
| 0% | 89.0% | 91.3% | +2.4 pp |
| 10% | 76.1% | 80.3% | +4.2 pp |
| 20% | 56.1% | 61.1% | +5.0 pp |
| 30% | 35.1% | 39.9% | +4.7 pp |
| 40% | 18.8% | 22.5% | +3.8 pp |
| All | 55.0% | 59.0% | +4.0 pp |

The lift is a ceiling, not an achieved rate: it assumes a user who inspects every alternative and always selects the faithful one. No user performed that selection in this benchmark.

#### eTable 8. Message-deduplicated primary corruption slope

Panel A. Original seven-model realized-corruption slope.

Sensitivity of the primary adjusted realized-corruption slope to input duplication. The matched controls (CTRL) are twins of AUTH items by message identity, not by corrupted-input bytes: of 91,700 control rows whose message-and-condition key matched an AUTH twin, only 19,838 (21.6%) were byte-identical to that twin, and almost all of those were at a target CER of 0, where zero realized errors force the corrupted input to equal the intended text. Every other control row used an independent corruption draw. The slope was refit on a count-balanced, message-deduplicated panel of the 2,299 unique messages (700 rows per message).

| Panel | Realized-corruption slope (log-odds) | 95% CI | Generations | Unique messages |
| --- | --- | --- | --- | --- |
| Full panel (adjusted primary) | 5.408 | 5.007 to 5.810 | 1,701,000 | 2,299 |
| Message-deduplicated | 5.587 | 5.226 to 5.949 | 1,609,300 | 2,299 |

Deduplication moved the slope by +0.179 log-odds (5.587 versus 5.408), so the primary estimate is not an artifact of repeated identical inputs.

Panel B. Current twenty-model target-CER primary slope.

The same message-identity duplication check, applied to the study's current primary benchmark question (target CER as the experimentally assigned exposure) on the current 20-model panel, not the original seven-model realized-CER slope in Panel A. As reported in Methods (Message corpora), CTRL's 131 message ids are a full subset of AUTH's 2,168 (CTRL items are matched controls drawn from the AUTH vocabulary), while CRIT's 131 items are a wholly separate set; dropping all CTRL-corpus rows removes this duplication and leaves the same 2,299 unique messages (2,168 AUTH + 131 CRIT) as Panel A. Unlike the original seven-model panel, where every model ran an identical replicate grid and dropping CTRL produced an exact 700-rows-per-message balance, the current 20-model panel includes three reduced-replicate reasoning models (gpt-oss:20b, gpt-oss:120b, deepseek-r1:32b), so the deduplicated panel is only near count-balanced: 1,751 rows for 1,525 messages (66.3%), 1,748 rows for 763 messages (33.2%), and 1,732-1,739 rows for the remaining 11 messages (0.5%).

| Panel | Target-CER odds ratio (per 10pp) | 95% CI | Generations | Unique messages |
| --- | --- | --- | --- | --- |
| Full panel (adjusted primary) | 2.296 | 2.275 to 2.317 | 4,252,326 | 2,299 |
| Message-deduplicated | 2.297 | 2.276 to 2.318 | 4,023,077 | 2,299 |

Deduplication moved the odds ratio by +0.001 (2.297 versus 2.296), so the current primary estimate is not an artifact of repeated message identities across corpora.

#### eTable 9. Rule-free (detector-removed) matched CRIT-versus-CTRL conditional logistic regression

Panel A. Original seven-model rule-free contrast.

The criticality contrast re-estimated with the five rule-based critical-substitution detectors (negation flip, numeral change, recipient change, urgency change, actionable omission) removed from the drift label, so drift was decided by the meaning channel alone (a bidirectional natural-language-inference contradiction, or a cosine similarity below threshold, gated by fluency). Because those five detectors also define the CRIT categories, this removes the circularity in a contrast that uses them to score the outcome. The matched design is otherwise identical to eTable 4.

| Specification | Critical-item OR | 95% CI | Discordant pairs | Matched observations | Drift share |
| --- | --- | --- | --- | --- | --- |
| Shipped (five detectors in label) | 1.16 | 1.13 to 1.19 | 27,812 | 55,624 | 32.0% |
| Rule-free (meaning channel only) | 1.10 | 1.07 to 1.13 | 27,237 | 54,474 | 31.1% |

Removing the detectors relabeled 4,917 rows out of the drift class and none into it, lowering the overall drift share from 32.0% to 31.1%. The critical-item excess survived, attenuated from OR 1.16 to OR 1.10, so it is not an artifact of the detectors shared between the challenge-set definition and the outcome label. The shipped run reproduced the original manuscript OR (1.155, rounded to 1.16).

Panel B. Twenty-model pooled and per-model rule-free matched odds ratio, and validation against Panel A. Before trusting the twenty-model extension, the same pipeline was subset to the original seven models: it reproduced the Panel A rule-free odds ratio (1.10 [95% CI, 1.07-1.13], 27,237 pairs, 54,474 observations) to within 0.03 odds-ratio units, confirming the extension before the twenty-model figure below is used.

Pooled (all 20 models): rule-free OR 1.095 (95% CI, 1.077 to 1.114), 68,602 matched pairs, 137,204 matched observations. The corresponding absolute-scale estimate, the matched risk difference, was 1.50 percentage points (95% CI, 1.27 to 1.71).

| Model | Rule-free OR | 95% CI | Matched pairs |
| --- | --- | --- | --- |
| Phi-3-mini | 0.99 | 0.93 to 1.05 | 4,783 |
| Qwen-3.5-27B | 1.01 | 0.94 to 1.09 | 3,751 |
| Qwen-2.5-7B | 1.02 | 0.96 to 1.09 | 3,879 |
| Gemma-4-E4B | 1.03 | 0.97 to 1.11 | 3,779 |
| Command-R | 1.04 | 0.97 to 1.11 | 4,080 |
| Gemma-2-9B | 1.06 | 0.99 to 1.14 | 3,972 |
| Gemma-3-27B | 1.09 | 1.01 to 1.17 | 3,711 |
| Phi-4-14B | 1.10 | 1.02 to 1.18 | 3,954 |
| Gemma-4-31B | 1.11 | 1.02 to 1.20 | 3,378 |
| Qwen-3.6-27B | 1.11 | 1.03 to 1.20 | 3,710 |
| Granite-3.3-8B | 1.12 | 1.05 to 1.20 | 4,162 |
| Phi-4-mini | 1.14 | 1.07 to 1.22 | 4,345 |
| Llama-3.1-8B | 1.15 | 1.08 to 1.22 | 4,595 |
| Mistral-Small-24B | 1.16 | 1.08 to 1.24 | 4,137 |
| Gemma-4-12B | 1.16 | 1.08 to 1.25 | 3,893 |
| Qwen-3.6-35B-A3B | 1.18 | 1.10 to 1.27 | 3,871 |
| Llama-4-Scout | 1.21 | 1.13 to 1.30 | 3,945 |

Every one of these 17 per-model estimates is a point estimate, a 95% CI, and a matched-pair count from its own model's matched pairs; six of seventeen (Phi-3-mini, Qwen-3.5-27B, Qwen-2.5-7B, Gemma-4-E4B, Command-R, Gemma-2-9B) have a CI that includes 1. The remaining three models (deepseek-r1:32b, gpt-oss:20b, gpt-oss:120b) do not appear in this table: their per-model conditional logistic regression failed to converge (an unnormalized covariance matrix could not be computed after the model dropped roughly 3,400-3,480 of their 3,668 matched pairs for having no within-pair outcome variance). This is a direct consequence of their reduced-replicate 40,500-exposure grid (eMethods S1): with only about 3 to 4 replicate corruptions per message-by-target-CER cell instead of 20, most of a model's own CRIT-CTRL matched pairs land on the same outcome on both sides of the match, leaving too little within-pair variance for a per-model fit. The pooled 20-model estimate above is unaffected because it still includes these three models' matched pairs (raising the total from 63,783 to 68,602 relative to the prior 16-model pooled figure); pooling across the full panel supplies enough aggregate variance even where three individual per-model fits cannot converge. Matching balance and match-quality diagnostics, shared with eTable 4, are reported in eTable 4, Panel C.

#### eTable 10. Original seven-model drift by realized character error rate, and uniform-grid versus operating-CER-reweighted pooled drift

Panel A gives the AUTH detected drift rate by realized (not target) character error rate, the curve summarized in the main text. Panel B contrasts the headline pooled rates, which average the five uniform target-CER conditions, with an operating-CER-reweighted sensitivity estimate (the fixed drift curve reweighted by empirical operating weights, not regenerated).

Panel A. AUTH detected drift by realized-CER bin.

| Realized CER | Detected drift (%) | Generations |
| --- | --- | --- |
| 0.00-0.05 | 1.5 | 401,128 |
| 0.05-0.10 | 8.2 | 137,606 |
| 0.10-0.15 | 16.2 | 143,003 |
| 0.15-0.20 | 26.2 | 158,466 |
| 0.20-0.25 | 38.5 | 147,952 |
| 0.25-0.30 | 50.3 | 124,222 |
| 0.30-0.35 | 58.3 | 120,841 |
| 0.35-0.40 | 65.8 | 111,321 |
| 0.40-0.45 | 71.3 | 69,118 |
| 0.45-0.50 | 71.4 | 59,423 |

Panel B. Uniform-grid pooled drift versus operating-CER-reweighted drift, by corpus.

| Corpus | Uniform-grid pooled drift (%) | Operating-CER-reweighted drift (%) |
| --- | --- | --- |
| AUTH | 30.9 | 28.8 |
| CRIT | 33.4 | 30.7 |
| CTRL | 31.2 | 29.3 |

The uniform-grid pooled rate equally weights five target-CER conditions (0, 10, 20, 30, and 40%), including a no-corruption condition and a 40% upper-bound stress condition, so it is a stress-test summary rather than an expected clinical event rate. The reweighted column holds the drift-versus-realized-CER curve fixed and reweights it by the real, selection-count-weighted operating-CER distribution of the 29 bigP3BCI ALS participants (weighted mean CER 0.2136). This reweighting is derived from the real per-participant selection-level CER, not from the simulated exposure's own realized CER (which is uniform by construction, so reweighting by it would be circular). It is an approximate, directionally-informative complement, not a precise incidence rate: an unweighted one-participant-one-vote reweighting instead gave 30.6% for AUTH, materially different from 28.8%, because the 29-participant weight histogram is lumpy.

#### eTable 11. Drift-specific confidence calibration and high-confidence miss rate, all 20 models

Drift-focused calibration companion to eTable 2 (eMethods S7), extended from the original seven-model diagnostic to all 20 models. The drift-specific AUROC is the discrimination of a low-confidence score (1 minus stated confidence) for the drift label against all non-drift outputs (faithful and degraded pooled); values above 0.5 mean lower stated confidence is associated with drift. Expected calibration error (ECE) is computed for that same drift-risk score against the observed drift indicator. The drift false-negative rate (FNR) at 0.9 is the fraction of true drift outputs a "flag when stated confidence is below 0.9" rule would miss. N missing confidence is the count of outputs with no usable verbalized confidence for that model, excluded from the AUROC/ECE/FNR columns but retained in the missingness audit below. Models are ordered by drift-specific AUROC; raw confidence is never pooled across models, so no pooled row is given here (the missingness audit is the one figure pooled across the panel, and is descriptive only).

| Model | Drift-specific AUROC | ECE (drift-risk) | Drift FNR at 0.9 | N missing confidence |
| --- | --- | --- | --- | --- |
| Gemma-4-31B | 0.910 | 0.107 | 0.182 | 2 |
| Qwen-3.5-27B | 0.861 | 0.147 | 0.132 | 0 |
| Gemma-4-12B | 0.848 | 0.215 | 0.470 | 23 |
| Phi-4-14B | 0.836 | 0.265 | 0.583 | 21 |
| Qwen-3.6-27B | 0.836 | 0.197 | 0.361 | 0 |
| GPT-OSS-120B | 0.829 | 0.096 | 0.037 | 15 |
| Qwen-3.6-35B-A3B | 0.828 | 0.185 | 0.192 | 1 |
| GPT-OSS-20B | 0.805 | 0.124 | 0.123 | 4,118 |
| Mistral-Small-24B | 0.784 | 0.219 | 0.204 | 2 |
| Llama-3.1-8B | 0.778 | 0.281 | 0.270 | 1,942 |
| Gemma-4-E4B | 0.773 | 0.150 | 0.345 | 1 |
| Gemma-2-9B | 0.757 | 0.255 | 0.678 | 3 |
| Llama-4-Scout | 0.735 | 0.115 | 0.009 | 50 |
| Granite-3.3-8B | 0.730 | 0.272 | 0.275 | 1,038 |
| Command-R | 0.730 | 0.192 | 0.431 | 15 |
| Gemma-3-27B | 0.714 | 0.218 | 0.800 | 3 |
| Phi-3-mini | 0.695 | 0.408 | 0.920 | 4,368 |
| DeepSeek-R1-32B | 0.695 | 0.191 | 0.336 | 3,328 |
| Phi-4-mini | 0.675 | 0.340 | 0.857 | 273 |
| Qwen-2.5-7B | 0.672 | 0.145 | 0.157 | 611 |

A confidence gate set at 0.9 would miss as little as 0.9% of true drift (Llama-4-Scout) and as much as 92.0% (Phi-3-mini). Drift-specific AUROC and the FNR-at-0.9 miss rate do not always rank models the same way: Llama-4-Scout has the lowest miss rate in the panel despite a below-median drift-specific AUROC (0.735, rank 13 of 20), because AUROC is a threshold-free ranking measure while the FNR is evaluated at one specific operating point. The three reduced-replicate-grid reasoning models diverge from each other here as elsewhere in the panel: GPT-OSS-120B has both a low ECE (0.096) and the second-lowest FNR-at-0.9 (3.7%) in the entire panel, GPT-OSS-20B is mid-pack, and DeepSeek-R1-32B has a below-median AUROC (0.695, tied for 17th of 20).

**Missingness audit. Missing or unparsable verbalized confidence was not at random with respect to drift. Across the full 20-model panel, 15,814 of 4,252,326 rows (0.37%) had no usable confidence; among those rows the drift rate was 67.9%, against a 31.6% drift rate across all scored and unscored rows pooled, roughly a two-fold enrichment. Missingness was concentrated in three models (Phi-3-mini, 4,368; GPT-OSS-20B, 4,118; DeepSeek-R1-32B, 3,328) that together account for 11,814 of the 15,814 missing-confidence rows; the remaining 17 models each contributed 1,942 or fewer, the largest being Llama-3.1-8B. This two-fold enrichment is smaller than the earlier 16-model panel's roughly three-fold figure because the four added models do not uniformly share the earlier pattern of missingness being drift-enriched: GPT-OSS-20B in particular has an FNR-at-0.9 among the lowest in the panel but its missing-confidence rows are, unusually, almost never drift, pulling the pooled enrichment down even as Phi-3-mini and DeepSeek-R1-32B reinforce it. Every per-model, per-CER, and per-critical-status row elsewhere in this manuscript and supplement reports the drift rate over all rows in that group (including missing-confidence ones), never over the scored subset alone, to avoid understating drift in the groups where confidence is least often recoverable.**

#### eTable 12. Per-model realized-corruption slopes (20 models), two-way cluster-robust inference, and original seven-model collinearity and mediation

Panel A reports the univariate realized-corruption slope (message-clustered cluster-robust logit, drift on realized CER) fit separately within each of the 20 models, extended from the original seven-model diagnostic. These per-model slopes and their descriptive min-max range and median are the primary cross-model summary, reported descriptively because the 20 checkpoints are a convenience sample of available open-weight models, not an inferential population from which pooling across models could generalize. Panel B reports two-way (message-by-model) cluster-robust inference on the same univariate specification, quantifying how much the message-only standard error understates uncertainty once within-model dependence is also taken into account. Panel C retains the original seven-model collinearity and mediation diagnostic (of that seven-model panel's own adjusted realized-CER slope, 5.408; distinct from the panel-wide adjusted slope reported in Results and Panel B), unchanged from the previous revision.

Panel A. Per-model univariate realized-corruption slope, all 20 models.

| Model | Slope (log-odds) | 95% CI | Generations |
| --- | --- | --- | --- |
| Gemma-4-31B | 10.781 | 10.553 to 11.010 | 243,000 |
| Phi-4-14B | 10.393 | 10.172 to 10.615 | 243,000 |
| Qwen-3.6-27B | 10.237 | 9.999 to 10.476 | 243,000 |
| Qwen-3.5-27B | 10.219 | 9.980 to 10.459 | 243,000 |
| Gemma-2-9B | 10.008 | 9.778 to 10.239 | 243,000 |
| Mistral-Small-24B | 9.637 | 9.408 to 9.865 | 243,000 |
| Gemma-4-12B | 9.636 | 9.409 to 9.863 | 243,000 |
| Gemma-3-27B | 9.033 | 8.836 to 9.230 | 243,000 |
| Qwen-3.6-35B-A3B | 8.942 | 8.730 to 9.153 | 243,000 |
| Phi-4-mini | 8.528 | 8.337 to 8.718 | 243,000 |
| Granite-3.3-8B | 8.435 | 8.197 to 8.673 | 243,000 |
| Phi-3-mini | 7.683 | 7.490 to 7.876 | 243,000 |
| Llama-3.1-8B | 7.454 | 7.219 to 7.689 | 243,000 |
| Command-R | 7.445 | 7.247 to 7.643 | 243,000 |
| Llama-4-Scout | 7.367 | 7.230 to 7.504 | 243,000 |
| GPT-OSS-120B | 6.802 | 6.587 to 7.017 | 40,500 |
| Gemma-4-E4B | 6.719 | 6.561 to 6.876 | 243,000 |
| DeepSeek-R1-32B | 5.113 | 4.899 to 5.326 | 40,500 |
| GPT-OSS-20B | 5.087 | 4.925 to 5.248 | 40,326 |
| Qwen-2.5-7B | 4.096 | 3.984 to 4.208 | 243,000 |
| Range (min-max), all 20 positive | 4.10 to 10.78 | -- | -- |
| Median | 8.48 | -- | -- |

The two reduced-replicate-grid gpt-oss checkpoints and deepseek-r1:32b are not the three lowest slopes in the panel (Qwen-2.5-7B and Gemma-4-E4B, both on the full 243,000-generation grid, sit below GPT-OSS-120B), so the panel's overall slope range and median are not being driven by the reduced-replicate models occupying one extreme; GPT-OSS-20B and DeepSeek-R1-32B do fall in the lower third of the panel, consistent with these two reasoning models' wider CIs reflecting genuine estimation uncertainty on a smaller grid rather than a systematically different slope.

Panel B. Two-way (message-by-model) cluster-robust inference on the univariate realized-CER slope (beta 8.037, all 20 models, n = 4,252,326; 2,299 message clusters, 20 model clusters, 45,971 message-by-model intersection clusters). This is a univariate specification (drift on realized CER alone, no covariates), distinct from the adjusted, message-clustered, panel-wide realized-CER slope reported in Results, and also distinct from the original seven-model adjusted slope (5.408) in Panel C below; none of these three should be conflated. The one-way (message-only) sandwich reproduces statsmodels' own default clustered standard error to within 0.02% relative difference, validating the sandwich algebra before the two-way extension is trusted.

| Standard-error source | SE | 95% CI on log-odds slope |
| --- | --- | --- |
| Message-only (one-way) cluster-robust | 0.071 | 7.90 to 8.18 |
| Two-way (message-by-model) cluster-robust | 0.429 | 7.20 to 8.88 |
| Model-level cluster bootstrap (20 clusters; flagged unstable) | 0.428 | 7.27 to 8.96 |

The two-way standard error is roughly six times the message-only standard error, so message-only clustering materially understated uncertainty; the direction and steep positive magnitude of the slope were unchanged. The model-level cluster bootstrap gives a comparably wide interval but is reported with an explicit instability caveat, because 20 model clusters is still fewer than the 30-50+ generally recommended for a trustworthy cluster bootstrap or a model-clustered standard error. A message-level aggregation sensitivity (one row per message, drift proportion on message-mean realized CER, binomial-weighted) gives a comparable but not identical ecological slope of 3.60 (95% CI, 3.42 to 3.78; 2,299 messages), lower because aggregation to the message level removes within-message row-level variation the row-level slope uses. The fully crossed message-by-model variational-Bayes mixed model was attempted again on this expanded panel and again did not converge at 4,252,326 rows, so the two-way sandwich above is the tractable substitute (eMethods S7).

Panel C. Collinearity and mediation of the adjusted primary model (original seven-model diagnostic, unchanged).

| Diagnostic | Value |
| --- | --- |
| Variance inflation factor, realized CER | 4.80 |
| Variance inflation factor, error count | 6.24 |
| Variance inflation factor, character length | 2.44 |
| Adjusted slope (direct effect, mediators retained) | 5.408 |
| No-mediators slope (total effect, corrupted-negation and numeral dropped) | 5.415 |

All three algebraically related corruption predictors had a variance inflation factor below 10, so the adjusted realized-CER coefficient is estimable. Dropping the two corruption-mediator flags to estimate the total rather than the direct effect left the slope essentially unchanged (5.415 versus 5.408).

#### eTable 13. Per-study decoder corruption, corruption-model allocation and structure, and held-out-participant matrix sensitivity

Panel A. Per-study decoder corruption and reweighted drift sensitivity (unchanged from the previous revision).

The confusion matrix was built from three bigP3BCI studies (F, L, and N); ALS status was re-derived directly from each recording's patient-identification header, and all 29 contributing participants were ALS-status, with zero non-ALS controls. Per-study empirical CER and a per-study drift sensitivity are shown; the drift sensitivity holds the AUTH drift-versus-realized-CER curve fixed and reweights it by each study's own selection-count-weighted operating-CER distribution.

| Study | ALS participants | Selections | Empirical CER | Reweighted AUTH drift (%) |
| --- | --- | --- | --- | --- |
| Study F | 10 | 1,067 | 0.221 | 25.9 |
| Study L | 11 | 990 | 0.161 | 24.5 |
| Study N | 8 | 480 | 0.306 | 44.4 |
| Pooled (all 29, all ALS) | 29 | 2,537 | 0.2136 | 28.8 |
| ALS-only | 29 | 2,537 | 0.2136 | 28.8 |

Because zero non-ALS participants exist among studies F, L, and N, the ALS-only reweighted estimate is mathematically identical to the pooled estimate (28.8%); this is the requested ALS-only sensitivity and is a null-difference finding by construction, not an approximation. The per-study reweighted estimates do differ (studies have different observed CERs), giving a per-study range of 24.5% to 44.4% around the pooled 28.8%.

Panel B. Corruption-budget allocation between substitution, insertion, and deletion. The corruption-injection procedure split each message's assigned character-error budget into an indel budget (10% of the character-error budget) and a substitution budget (the remainder); within the indel budget a structural 50/50 coin flip in the code, not a free parameter chosen after the fact, decides deletion versus insertion.

| Corruption type | Fraction of the character-error budget |
| --- | --- |
| Substitution | 0.90 |
| Deletion | 0.05 |
| Insertion | 0.05 |

Out-of-grid characters (ordinary punctuation, outside the 36-symbol grid) can never be substituted, but are not otherwise protected: the indel branch fires before any grid-membership check, so an out-of-grid character can still be deleted or preceded by an inserted grid character. In the cached AUTH message sample, 3.57% of characters are structurally outside the grid (corruption-draw-independent); empirically, 3.19% of characters in the cached exposure file (50,480 rows checked) were recorded as passing through completely untouched, slightly below the structural figure as expected, since some out-of-grid characters are still touched by the indel branch.

Panel C. Row/column structure of the empirical confusion matrix, against a structure-free (uniform) null. The per-character substitution step draws the replacement from the intended character's own row of the empirical 36-by-36 confusion matrix (eFigure 2); this panel checks whether that matrix's off-diagonal mass is enriched for same-row/same-column grid neighbors relative to what a uniform, geometry-free substitution rule would produce.

| Off-diagonal mass | Empirical | Uniform-null expectation |
| --- | --- | --- |
| Same row as intended character | 18.6% | 14.3% |
| Same column as intended character | 23.5% | 14.3% |
| Neither same row nor same column | 58.0% | 71.4% |

Both same-row and same-column mass are enriched above the uniform-null expectation, consistent with the row-and-column flash sequence of P300 spelling: substitution already encodes real decoder structure, rather than being geometry-agnostic. This says nothing about independence across character positions within a message: the corruption code draws a fresh, uncorrelated outcome at every position, with no mechanism for correlated runs of errors that a phenomenon such as attentional fatigue could plausibly produce; that remains a genuine simplifying assumption, stated as a limitation rather than resolved here.

Panel D. Held-out-participant confusion-matrix sensitivity. This is a descriptive bound, not a full regeneration of the study: the unmodified corruption function was reapplied to the same cached AUTH messages (500 messages, five replicates per target CER), once per held-out participant's own confusion matrix and once with the cohort-pooled matrix used throughout, at matched target CER levels.

| Target CER | Pooled-matrix mean realized CER | Held-out-participant mean realized CER | Max absolute difference (pp) | Max relative difference |
| --- | --- | --- | --- | --- |
| 10% | 0.0957 | 0.0970 | 0.37 | 3.9% |
| 20% | 0.1916 | 0.1932 | 0.54 | 2.8% |
| 30% | 0.2890 | 0.2893 | 0.50 | 1.7% |
| 40% | 0.3881 | 0.3855 | 0.70 | 1.8% |

Across all four target-CER levels and 29 held-out participants, the realized CER a single participant's own confusion matrix would have produced diverges from the cohort-pooled matrix by at most 0.7 percentage points, under 4% relative. This bounds, without eliminating, the possibility that a different confusion matrix would materially change the study's exposure; a full per-participant regeneration of the labeled cohort was out of scope for this revision (it would require new model inference).

#### eTable 14. Model manifest for all 20 models: served tag, quantization, ollama config digest, immutable model-layer digest, and hardware/configuration confirmation

A reproducible manifest for all 20 models named in Methods ("Models and prompting condition"). The seven rows for the original panel are reused verbatim from the earlier reviewer-requested manifest, assembled from local run artifacts and, for the ollama registry config digest, fetched directly from the ollama manifest store (local scratch storage). The nine rows added for the first panel-expansion pass (command-r:latest, gemma2:9b, gemma3:27b, llama3.1:8b, llama4:17b-scout-16e-instruct-q4_K_M, phi3:mini, qwen2.5:7b, qwen3.6:27b, qwen3.6:35b-a3b) originally had no local model-configuration capture, so an earlier version of this table reported their ollama config digest and quantization as not recorded. Both were subsequently recovered for these nine by reading directly from the ollama manifest store (read 2026-08-03): the seven config digests that overlap the earlier capture match it byte-for-byte, and three previously null tags (qwen3.6:27b, gemma3:27b, phi3:mini) were independently re-read from the manifest store and also matched byte-for-byte, so the recovery reproduces rather than replaces the original values, and nothing below is inferred or guessed. A final four rows (granite3.3:8b, gpt-oss:20b, gpt-oss:120b, deepseek-r1:32b) complete the panel at 20 models; their digests were recovered by the identical manifest-store read (read 2026-08-11). The table also reports each model's immutable model-layer digest, the sha256 of the served weights layer itself rather than the smaller config layer, so that every one of the 20 served checkpoints is exactly identifiable regardless of any later change to a mutable tag. Inference-library version, chat-template identifier, and context-window size remain not recorded for all 20 models: the context-window size was never set in the runner or any submission script, so Ollama's server-side default applied but was not independently confirmed against the server install, and the inference-library (Ollama) version and chat-template identifier were never captured by the run's provenance logging. Parameter count for the thirteen expansion models is reported only when a numeric token is literally present in the served tag, not imported from an outside model card; this includes the two gpt-oss checkpoints, whose served tags carry no mixture-of-experts token analogous to llama4's "16e", so no active-parameter or expert-count claim is made for them beyond the literal total-parameter token in the tag. Every model, across all 20, was served at temperature 0 against the identical frozen prompt bank named in Methods (SHA-256 97d7a1f9f8dc7fba731537028a1271ab8f53fecdeb21bcd927378524ed0f7696 for all 20). The 17 non-reasoning models were served with think-mode off and an output-token limit of 160; the two gpt-oss checkpoints and deepseek-r1:32b interleave chain-of-thought reasoning before the final answer by design and were instead run at a larger output-token limit sized to accommodate that reasoning trace (eMethods S1). All models ran on a SLURM-managed GPU compute cluster, 1x NVIDIA L40S GPU (48GB VRAM) per task. The rightmost column reports whether a surviving on-disk artifact directly names that model's served tag together with this hardware and run configuration: for the original seven models and two of the first nine expansion models (command-r:latest, llama3.1:8b), this is a per-model SLURM submission script. For the final four models the submission script itself is a shared, parameterized launcher (the served tag is passed at submission time via an environment variable, not hardcoded in the script text), but each array task's own surviving stdout log directly prints its served tag together with hostname and GPU device index at run start, which this table treats as an equally direct, non-inferred confirmation and flags "Yes" accordingly. For the remaining seven of the first nine expansion models the same hardware and run configuration are inferred rather than directly confirmed, from a shared per-model output-file naming pattern their 243,000-row production data shares with the two directly-confirmed models and from this manuscript's own stated model list, not from a surviving per-model submission script or run log, and are flagged "No" rather than presented as equally confirmed.

| Served tag | Parameters | Quantization | Ollama config digest (sha256, first 12) | Model-layer digest (sha256, first 12) | Hardware + config independently confirmed |
| --- | --- | --- | --- | --- | --- |
| gemma4:e4b | 7.5B | q8 (Q8_0) | ce677f641308 | 0213bca357a6 | Yes |
| gemma4:12b | 11.9B | q8 (Q8_0) | a298f5e22b42 | 6519da92d5ed | Yes |
| gemma4:31b | 30.7B | q8 (Q8_0) | 8e73751606f4 | 66fda95ed640 | Yes |
| phi4:mini | 3.8B | q8 (Q8_0) | cc7678ebeb6c | 7c6dee0881c1 | Yes |
| phi4:14b | 14.7B | q8 (Q8_0) | 9a8b7f7ee4ca | f68df5d3a22a | Yes |
| qwen3.5:27b-q4 | 26.9B | q4 (Q4_K_M) | 74433bac6fee | 355d2d2f3209 | Yes |
| mistral-small:24b | 23.6B | q4 (Q4_K_M) | a4ae56844628 | 5e84c54ee8d5 | Yes |
| command-r:latest | not recorded | q4 (Q4_0) | 8e63f21e12fb | 8e0609b8f0fe | Yes |
| gemma2:9b | ~9B (from served tag) | q4 (Q4_0) | 10aa81da732e | ff1d1fc78170 | No |
| gemma3:27b | ~27B (from served tag) | q4 (Q4_K_M) | f838f048d368 | e796792eba26 | No |
| llama3.1:8b | ~8B (from served tag) | q4 (Q4_K_M) | 455f34728c9b | 667b0c1932bc | Yes |
| llama4:17b-scout-16e-instruct-q4_K_M | ~17B active, 16-expert MoE (from served tag) | q4 (Q4_K_M) | f7ce8f326f5d | 9d507a36062c | No |
| phi3:mini | not recorded | q4 (Q4_0) | 23291dc44752 | 633fc5be925f | No |
| qwen2.5:7b | ~7B (from served tag) | q4 (Q4_K_M) | 2f15b3218f05 | 2bada8a74506 | No |
| qwen3.6:27b | ~27B (from served tag) | q4 (Q4_K_M) | 728c795c7762 | 83c54730a5fe | No |
| qwen3.6:35b-a3b | ~35B total, ~3B active MoE (from served tag) | q4 (Q4_K_M) | 5d1c86a949f7 | f5ee307a2982 | No |
| granite3.3:8b | ~8B (from served tag) | q4 (Q4_K_M) | 122661774644 | 77bcee066a76 | Yes |
| gpt-oss:20b | ~20B (from served tag) | q4 (MXFP4) | 776beb3adb23 | e7b273f96360 | Yes |
| gpt-oss:120b | ~120B (from served tag) | q4 (MXFP4) | 0b3eaefc220f | 6be6d66a3f54 | Yes |
| deepseek-r1:32b | ~32B (from served tag) | q4 (Q4_K_M) | c7f3ea903b50 | 6150cb382311 | Yes |

Full 64-character digests and prompt-bank hashes, and the per-model source notes behind every "not recorded" and "Yes"/"No" entry above, are in the underlying reproducibility archive. Quantization and the model-layer digest are now confirmed for all 20 models, read directly from the ollama manifest store rather than inferred from the served tag: Q8_0 (8-bit) for the three gemma4 checkpoints and both phi4 checkpoints (5 models), Q4_K_M (4-bit) for qwen3.5:27b-q4, mistral-small:24b, gemma3:27b, llama3.1:8b, llama4:17b-scout-16e-instruct-q4_K_M, qwen2.5:7b, qwen3.6:27b, qwen3.6:35b-a3b, granite3.3:8b, and deepseek-r1:32b (10 models), Q4_0 (4-bit) for command-r:latest, gemma2:9b, and phi3:mini (3 models), and MXFP4 (a native 4-bit microscaling format) for the two gpt-oss checkpoints (2 models). Only qwen3.5:27b-q4's and llama4:17b-scout-16e-instruct-q4_K_M's served tags spell their quantization out explicitly, so before this recovery the served tag alone was an incomplete guide to numeric precision across the panel; the manifest-store read resolves that gap for every model rather than only these two self-labeled tags. Because the panel still spans differing parameter counts and quantizations, and, for seven of the thirteen expansion models, an unconfirmed exact serving configuration beyond model identity itself, any ranking of models by drift rate, calibration, or any other outcome elsewhere in this manuscript remains descriptive variation across checkpoints as actually served, not a controlled comparison of model quality, and does not isolate the effect of model identity from these confounded serving differences: the immutable digests in this table identify exactly which checkpoint produced each result, but do not by themselves equalize quantization or serving configuration across the panel.

#### eTable 15. LLM-BCI reporting checklist crosswalk (Gorenshtein et al, ref 4)

Compliance crosswalk against the reporting checklist proposed in our earlier systematic review (ref 4, Section 4.3), item-by-item. That checklist scopes its scenario-specific items by study design; this in-silico benchmark is an offline LM benchmark with no participant, device, or live decoder loop (Methods, "Study design and reporting"), so Panel A (items required of every study regardless of design) and Panel B (the offline-LM-benchmark scenario) apply, and Panel C records the scenario-specific items ref 4 defines for designs this study is not: live copy-spelling, intent-based/dialogue, and command-and-control/passive-affective. An item is marked "Not reported" when it was in scope but not captured, and "Not applicable" when the study design places it out of scope; both are distinguished from full compliance rather than folded into a single pass/fail.

Panel A. Items required of every study, regardless of design

| Checklist item (ref 4, Section 4.3) | Status | Where reported |
| --- | --- | --- |
| (a) Hardware montage: device model, channel count, channel labels, sampling rate, reference, ground and impedance handling | Not applicable -- no participant or device was used in this study; the montage of the source decoding studies (36-symbol BCI2000 grid; bigP3BCI Studies F, L, N) is reported for those studies, not re-reported here | eTable 13; refs 14, 15 |
| (b) Integration depth: decoder, interface or dialogue | Reported -- interface layer (post-editing of decoder output) | Introduction ¶1; Methods, "Message corpora" |
| (c) Study-design category: online real-time, offline simulation or system proposal | Reported -- offline simulation | Methods, "Study design and reporting" |
| (d) LLM identifier and version, API endpoint or local deployment, prompt template and decoding settings (temperature, top-p, max tokens) | Partially reported -- served tag, quantization, and digests per model; local deployment; frozen six-prompt bank fixed by SHA-256 hash; temperature (0) and output-token limit reported; top-p not recorded | Methods, "Models and prompting condition"; eTable 14; eMethods S1 |
| (e) Latency breakdown: biological dwell time, decoder execution time, LLM query-to-response time | Not reported -- an offline batch benchmark has no live user, so biological dwell time and decoder execution time do not apply; LLM query-to-response latency was not captured by run provenance logging | -- |
| (f) Ablation comparators: at minimum a no-LM baseline and a classical n-gram baseline on the same data | Not reported -- no run compared the LLM-corrected output against sending the raw corrupted text uncorrected, or against a classical n-gram baseline | -- |

Panel B. Offline LM benchmark scenario (the applicable scenario for this study)

| Checklist item (ref 4, Section 4.3) | Status | Where reported |
| --- | --- | --- |
| MRR@k and Recall@k at k ∈ {3, 5, 10} | Not applicable -- the outcome is a categorical faithful/degraded/drift adjudication of a full free-text reconstruction (Methods; eMethods S4), not a ranked-retrieval task, so MRR/Recall@k have no defined analogue here | Methods, "Outcome adjudication" |
| Performance under injected noise that matches the target BCI character-error rate | Reported -- the primary exposure is synthetic corruption at five target CER levels (0%-40%) from an empirical P300 confusion matrix | Methods, "Corruption model"; Figure 1a; eTable 3 |
| A clear statement that any reported typing-speed number is theoretical until the LM is coupled to a real decoder | Not applicable -- no typing-speed, information-transfer-rate, or keystroke-savings number is reported anywhere in this manuscript | -- |

Panel C. Scenario-specific items not applicable to this study's design

| Checklist scenario (ref 4, Section 4.3) | Status |
| --- | --- |
| Copy-spelling scenario: selection and character accuracy, Wolpaw information transfer rate, characters per minute, time to complete a standardized corpus, keystroke savings against an equivalent no-LM speller | Not applicable -- no live decoder loop or completed spelling session; per ref 4, information transfer rate is appropriate only to a copy-spelling scenario with a real decoder |
| Intent-based or dialogue scenario: semantic accuracy, semantic information transfer rate, task-goal success rate per scenario per subject, LLM selections per completed task | Not applicable -- no dialogue or task-completion design |
| Command-and-control or passive affective scenario: command decoding accuracy, emotion-classification accuracy, end-to-end task success, explicit statement that information transfer rate and keystroke savings do not apply | Not applicable -- not a command or passive-affective BCI design |

### eFigures

#### eFigure 1. Realized versus target character error rate

Mean realized character error rate (points) against the five target levels (0, 10, 20, 30, and 40 percent). Error bars show plus or minus one standard deviation of the realized character error rate across every corrupted draw at that target level, that is, across all intended messages, each crossed with 20 independently seeded replicate corruptions (the population that was carried forward as the analytic exposure); the target-0 condition returns the text unchanged, so its realized rate is exactly 0 with no spread. Realized values approximated their targets across the range (for example, a 20 percent target realized approximately 17.5 percent, and a 40 percent target realized approximately 37.6 percent). The spread reflects that a fixed target rate produces variable realized corruption across messages of different lengths and across replicate draws.

#### eFigure 2. Pooled 36-by-36 P300 character confusion matrix

Heatmap of the probability of the selected symbol conditional on the intended symbol for the 36-symbol BCI2000 speller grid (A through Z, the digits 1 through 9, and space, the 36th symbol, labeled SP on both axes), pooled across 29 subjects, 2,537 selections, and 425 files from bigP3BCI studies F, L, and N. Rows sum to 1; the diagonal is the correct selection. Off-diagonal mass concentrates within the same speller row or column, consistent with the row-and-column flash sequence of P300 spelling.

#### eFigure 3. Per-model confidence-reliability small multiples

Confidence-reliability curves for each of the 20 evaluated models individually: mean stated confidence (10 equal-width bins) plotted against the observed faithful rate, against the diagonal of perfect calibration, with each model's own expected calibration error (ECE) and AUROC printed in its panel. A muted pooled curve is shown in every panel for visual context only. Verbalized model confidence is not calibrated on a common scale across models (Methods), so these per-model curves, not a single pooled curve, are the primary display of calibration heterogeneity here; the per-model ECE and AUROC are also shown as the main-text Figure 3, and the pooled meta-analytic ECE and AUROC (random-effects, DerSimonian-Laird, with 95% CI), reported as a descriptive cross-model summary rather than a shared-scale claim, are in eFigure 4.

This figure was moved out of the main figure sequence, where an earlier version of Figure 1 embedded it as a third panel, because a 20-panel grid is not legible at main-text print size. No computation changed in the move: every curve, bin, and printed value is identical to the panel it replaced.

#### eFigure 4. Pooled confidence-outcome calibration (descriptive)

(a) Pooled confidence-outcome curve, all 20 models combined: mean stated confidence (10 equal-width bins) plotted against the observed faithful rate, against the diagonal of perfect calibration. The shaded gap marks bins where stated confidence exceeded the observed faithful rate, and marker area scales with the log of each bin's count. Because verbalized confidence is not on a common scale across models (Methods), this pooled curve is a descriptive cross-model summary and is not interpretable as calibration for any individual model, as the in-panel note states; the per-model reliability curves in eFigure 3 are the interpretable per-model display.

(b) Pooled meta-analytic expected calibration error (ECE 0.32, 95% CI 0.27-0.37) and AUROC (0.83, 95% CI 0.80-0.85) for confidence discriminating faithful from non-faithful (degraded or drift) outputs, random-effects (DerSimonian-Laird) pooling across all 20 models, with between-model heterogeneity (I^2^, exceeding 99% for both metrics) printed alongside each pooled value.

This pooled display was moved out of the main figure sequence in revision, where it briefly stood as Figure 3; the interpretable per-model ECE and AUROC that discriminate faithful from non-faithful outputs are now the main-text Figure 3. No computation changed in the move: every bin, curve, and pooled value is identical.

### Code and data availability

The intended-message corpora are the Costello/Boston Children's Hospital ALS message-banking vocabulary and a message-critical probe set; corpora are cited and raw phrase lists are not redistributed. The decoder-noise anchor is derived from the openly available bigP3BCI dataset. eTable 1 gives a dated timeline of the study's development. Analysis code, the frozen prompt bank, the frozen exposure file, and the derived result digests are archived at <https://github.com/BRIDGE-GenAI-Lab/BCI-Intent-drift-> and, as a public archive, on the Open Science Framework. The sixteen-model physician panel described in eMethods S6 is the primary measurement-validation analysis: two physicians rated the full 2,281-item instrument (16 of the study's 20 models, all 3 corpora, the full CER grid; it predates the four models added last, eMethods S1), a third physician (O.D.) adjudicated their 343 disagreements, and inter-rater agreement, ensemble-versus-physician-consensus validation, and the weighted misclassification correction are reported in eMethods S6 and eTable 6 Panel E (Cohen kappa 0.76 and 0.41, respectively; corrected AUTH drift 27.9%). The earlier seven-model physician panel described in eMethods S6 is reported as a sensitivity analysis; its class-specific agreement with the automated ensemble and its misclassification-corrected drift estimate are in eTable 6 Panels A-D. Requests for further detail may be directed to the corresponding author.
